# A *de novo* human *MYCBP2* variant enhances memory persistence across species

**DOI:** 10.1101/2024.12.18.24319147

**Authors:** Andreas Papassotiropoulos, Dmytro Nesterenko, Daniel Beck, Desirée Böck, Jana Petrovska, Andreas Arnold, Aurora K.R. LePort, Virginie Freytag, Nathalie Schicktanz, David Coynel, Aline Baur, Claudia Riva, Melissa Long, Pavlina Mastrandreas, Melanie Neutzner, Vanja Vukojevic, Attila Stetak, Noëlle Burri, Vaibhav Gharat, Navid Ghaffari, Janani Durairaj, Susanne Mueller, Torsten Schwede, Oliver Bieri, Johannes Gräff, Efthimios M.C. Skoulakis, Katharina Henke, Bettina Burger, Sven Cichon, Philipp Boehm-Sturm, Pawel Pelczar, Verdon Taylor, Craig E.L. Stark, James L. McGaugh, Gerald Schwank, Johannes Bohacek, Camin Dean, Dominique J.-F. de Quervain

**Author notes:** Corresponding authors: Andreas Papassotiropoulos: Division of Molecular Neuroscience, Department of Biomedicine, University of Basel, Birmannsgasse 8, CH-4055 Basel, Switzerland, Dominique J.-F. de Quervain: Division of Cognitive Neuroscience, Department of Biomedicine, University of Basel, Birmannsgasse 8, CH-4055 Basel, Switzerland, Camin Dean: German Center for Neurodegenerative Diseases, Charité University of Medicine, 10117 Berlin, Germany, Johannes Bohacek: Laboratory of Molecular and Behavioral Neuroscience, Institute for Neuroscience, Department of Health Sciences and Technology, ETH Zurich, 8092 Zurich, Switzerland, Gerald Schwank: Institute of Pharmacology and Toxicology, University of Zurich, 8057 Zurich, Switzerland. These authors contributed equally to the study (Dmytro Nesterenko, Daniel Beck, Desirée Böck, Jana Petrovska, Andreas Arnold).

## Abstract

Exceptional human memory phenotypes provide an entry point to identify rare coding variants that influence memory capacity. Here we studied an individual with highly superior autobiographical memory (HSAM), a rare condition characterized by the exceptional ability to recall personal events across decades with striking detail. The individual showed extraordinary autobiographical recall despite otherwise average cognitive performance. Whole-brain structural imaging revealed exceptionally large volumes in hippocampal subregions previously linked to autobiographical memory. Trio whole-exome sequencing identified a rare *de novo MYCBP2* V768F missense variant, NM_015057.5:c.2302G>T, p.(Val768Phe) (hereafter V768F), absent from more than 700,000 population exomes and affecting an evolutionarily conserved residue in the encoded E3 ubiquitin-protein ligase. To test the functional impact of this human variant, we engineered the homologous substitution in *C. elegans* and mice. *C. elegans* carrying the homologous variant showed enhanced long-term associative memory without detectable sensory, motor, or growth changes. Heterozygous Mycbp2 knock-in mice carrying the orthologous substitution showed improved long-term memory retention in spatial and non-spatial memory tasks. Finally, prime editing of the same variant selectively in dorsal hippocampal neurons of adult mice improved spatial long-term memory, indicating that this effect does not require developmental expression. These findings demonstrate that a rare *de novo MYCBP2* variant identified in an individual with HSAM enhances long-term memory retention when the homologous substitution is introduced into *C. elegans* and mice.

## Background

Highly Superior Autobiographical Memory (HSAM) is an exceptionally rare condition, with approximately 100 individuals identified so far^1–3^. Notwithstanding reports dating back to the 19^th^ century^4^, HSAM first gained attention in 2006 through the case study of “AJ,” a woman who could remember autobiographical events of almost every day of her life since adolescence. That report showcased a form of memory performance strikingly different from typical or even other exceptional memory types, such as those observed in savants^5^.

Studies show that HSAM and non-HSAM individuals recall a similar number of autobiographical details from the preceding week, indicating comparable encoding of events^6^. However, while recall over extended intervals, spanning the past month, year, and decade, severely declines in people without HSAM, it shows only minimal decay over time in HSAM individuals^6^. This unique trait suggests that HSAM may be driven less by an enhanced capacity to learn and more by sustained memory retention^1, 7, 8^.

Exceptional autobiographical memory is shared among all HSAM individuals; however, there is heterogeneity in their overall cognitive profiles. Not all exhibit the same memory strength across different types or contents of memory, and variations exist in their cognitive, clinical, and neural profiles^7, 8^. This diversity suggests that multiple, hitherto unknown individual biological mechanisms might underlie HSAM.

Here we report the case of an individual with HSAM who underwent neuropsychological testing and structural brain imaging. Whole exome sequencing was performed in this individual and their parents to assess whether a *de novo* genetic variant might underlie the exceptional autobiographical memory, given that neither parent shared the phenotype. To test the causal effect of the identified *de novo* variant on memory persistence, we introduced the homologous variant into the germline of *C. elegans* and mice using CRISPR-Cas9, and postnatally introduced the variant selectively in dorsal hippocampal neurons of adult mice via prime editing.

## Results

### Neuropsychological assessment of the HSAM participant

The male proband, herein referred to as JN, was previously identified as exhibiting highly superior autobiographical memory (HSAM)^6, 7^ and reports being right-handed. We invited JN, now in his sixties, to our research institute to obtain a detailed cognitive profile and a structural brain scan. The study protocol was approved by the Ethics Committee of Northwest and Central Switzerland.

We validated JN’s HSAM status through the administration of the 10 Dates Quiz^9^, which assesses autobiographical memory by asking participants to recall the day of the week, a verifiable public event that occurred within ± one month of the generated date, and a personal event for ten randomly selected dates, ranging from age fifteen to the present. JN correctly identified the day of the week for all ten dates (10/10), recalled a verifiable public event for nine dates (9/10), and reported a personal autobiographical event for all ten dates (10/10), resulting in a total score of 29 out of 30. A score of 20 points or higher qualifies an individual for HSAM, whereas non-HSAM controls typically score below 3 points^9^.

To characterize JN’s broader cognitive profile, we administered standardized, age-matched neuropsychological tests covering general intellectual ability, verbal and non-verbal memory, attention, and processing speed (Fig. 1a; Supplementary Table 1). Performance was largely within the average range across measures. Full-scale IQ was average (WAIS-IV FSIQ = 104), as were indices of verbal comprehension, perceptual reasoning, processing speed, auditory memory, visual memory, immediate memory, and delayed memory. Notably, the WMS-IV Delayed Memory Index assesses retention after a delay of approximately 20–30 min rather than over days or longer. Working memory fell in the high-average range, whereas one Stroop subtest was in the borderline below-average range. Together, these findings indicate that JN’s broader cognitive profile was consistent with a selective enhancement of autobiographical memory.

**Fig. 1.**
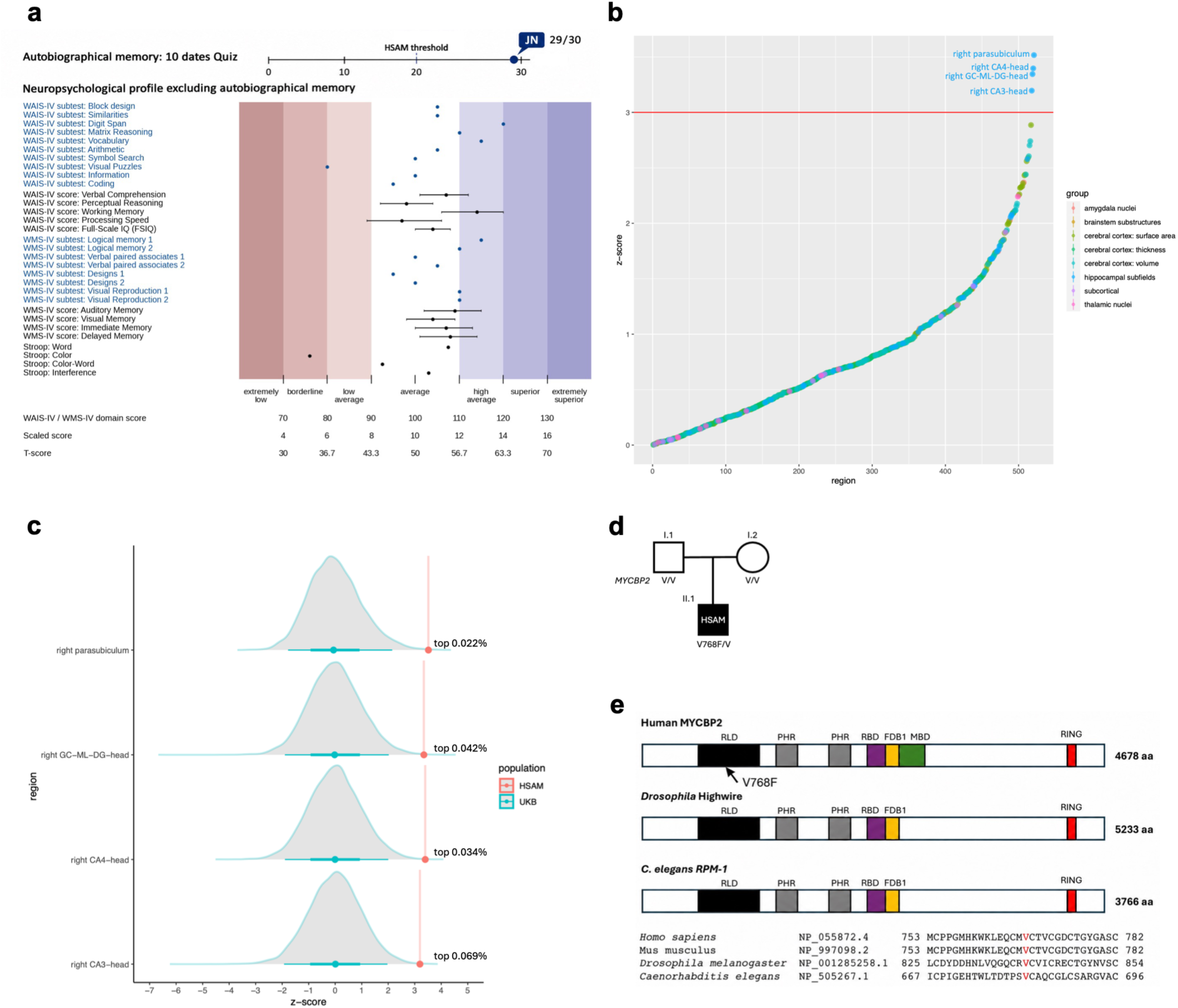
MYCBP2 variant in the individual with HSAM. **a**, JN’s neurocognitive profile. Blue dots represent 10 dates Quiz score, and individual WAIS-IV and WMS-IV subtest scores; black dots represent composite scores and Stroop test scores. Black horizontal whiskers indicate reference 95% confidence intervals for the composite scores. **b,** Comparison of JN’s image-derived phenotypes of cortical and subcortical brain regions to a normative dataset from the UK Biobank. Dots represent positive Z values in ascending order. Dots above the red horizontal line exceed Z = 3, indicating extremely large values of the corresponding regions in JN as related to the reference population. GC-ML-DG: granule cell and molecular layers of the dentate gyrus. **c,** Distribution of the volumes of the four brain regions exceeding Z = 3 in the reference population. Red dots and vertical lines denote JN’s volumes in these regions. **d,** *MYCBP2 de novo* variant in the family trio. **e,** Conservation of protein domains (RCC1-like GEF domain (RLD), PHR family specific domains (PHR), RAE-1 binding domain (RBD), FSN-1 binding domain 1 (FBD1), Myc binding domain (MBD) and RING-H2 ubiquitin ligase domain (RING)) and of the mutated residue. The sequence alignment shows a 30-residue window corresponding to human MYCBP2 residues 753–782. Relative to the human sequence, amino acid identity is 100% (30/30) in *Mus musculus*, 43.3% (13/30) in *Drosophila melanogaster* and 40.0% (12/30) in *C. elegans*. Sequence similarity, defined as identical residues plus non-identical substitutions with BLOSUM62 scores >0, is 100%, 53.3% and 53.3%, respectively. The valine corresponding to human V768 is identical in all four species.

### Structural brain imaging

We next investigated whether JN showed distinctive structural brain features compared to a normative dataset. This comparison utilized the UK Biobank database to assemble a comparison sample of 7,755 men aged 58-68 (see Methods). We employed the UK Biobank’s acquisition protocol and pipeline for structural imaging, yielding image-derived phenotypes (IDPs) related to cortical volume, thickness, and surface area, as well as volumes of subcortical regions. To quantitatively assess JN’s brain structure, his ranking within the distribution of each IDP among the control group was calculated through standardization of his raw measurements, with Z > 3, denoting extremely large values^10^.

Brain volumetry based on FreeSurfer’s automatic segmentation^11–14^ showed that the volumes of the right parasubiculum (Z = 3.52, corresponding to top 0.022% of the reference population), right head of the granule cell and molecular layers of the dentate gyrus (Z = 3.35, i.e., top 0.04%), right head of the CA4 region (Z = 3.40, i.e., top 0.034%), and right head of the CA3 region (Z = 3.20, i.e., top 0.07%) exceeded the Z = 3 threshold, indicating extremely large volumes of these regions of the hippocampal formation in JN compared to the reference population (Fig. 1b,c; Supplementary Fig. 1). The corresponding left-sided subfields also had positive Z-scores (1.75–2.58) but did not exceed the predefined Z = 3 threshold (Supplementary Table 2). No other subcortical IDPs exhibited volumes exceeding Z = 3 (Supplementary Table 2). Likewise, none of the IDPs for cortical volume, cortical thickness, or cortical surface area showed unusually large values (Z > 3; Supplementary Table 2). In summary, exclusively regions within the hippocampal formation, areas previously linked to autobiographical memory^15–19^, exhibited exceptionally large volumes in this individual with HSAM. Of note, among the 7755 UK Biobank participants, none demonstrated volumes exceeding a Z-score of 3 in all four sub-regions of the hippocampal formation, a feature uniquely present in JN.

### Detection of de novo variants by whole exome sequencing

According to JN’s family history, neither his parents nor his two siblings (one brother and one sister) had memory abilities comparable to his own. None of these relatives had undergone formal HSAM assessment, and JN has no children. Given the absence of reported HSAM in his non-consanguineous parents, we hypothesized that a *de novo* variant might contribute to his exceptional memory. To test this, we performed whole exome sequencing on the participant and his parents.

Variants identified as *de novo* by both the GATK refinement workflow and VarScan2 were considered high-confidence candidates. For functional follow-up, we applied predefined prioritization criteria of absence from the respective Genome Aggregation Database gnomAD v2.1.1 (encompassing over 125,000 individual exomes) given the rarity of HSAM, and high *in silico* probability of functional impact (i.e., Combined Annotation Dependent Depletion (CADD) score >20). Both calling pipelines identified four high-confidence *de novo* variants: missense variants in *MYCBP2* and *DIS3L*, a non-coding indel annotated to *FPGT/FPGT-TNNI3K/TNNI3K*, and an intronic variant in *LMCD1*. Comparative assessment of the four high-confidence *de novo* candidates integrated gene-level constraint and biological evidence in addition to population frequency and predicted functional impact (Supplementary Table 3). The *DIS3L* missense variant had a higher CADD PHRED score than *MYCBP2* V768F (31 versus 24.2), but was present in gnomAD v2.1.1 at allele frequency of 2.9 × 10⁻⁴, whereas *MYCBP2* V768F was not observed. *MYCBP2* also showed substantially stronger constraint against predicted loss-of-function and missense variation than *DIS3L*. Biologically, *MYCBP2*, which encodes an E3 ubiquitin-protein ligase and is a member of the PHR (Phr1/MYCBP2, highwire and RPM-1) family of proteins^20–22^, has established roles in axon development and synaptic function^20, 23–25^. Disease-associated *MYCBP2* variants further demonstrate its importance for nervous system function^26^, although these findings do not establish the functional direction of V768F. *DIS3L*, by contrast, participates in cytoplasmic RNA turnover and quality control and is expressed in the brain, but has no established specific role in memory or synaptic plasticity. The remaining candidates (i.e., the non-coding indel annotated to *FPGT/FPGT-TNNI3K/TNNI3K* and the intronic *LMCD1* variant) were also absent from gnomAD, but had substantially lower CADD scores (1.3 and 6.5, respectively). *FPGT* functions in fucose salvage, *TNNI3K* encodes a predominantly cardiac kinase, and *LMCD1* is a transcriptional cofactor. Although brain-related observations have been reported for these genes, including reduced *FPGT* transcript levels in human Alzheimer’s disease brain tissue and 5xFAD mice^27^ and *Lmcd1* expression in embryonic mouse midbrain progenitors^28^, these expression findings do not by themselves establish a direct role of either gene in memory or a functional consequence of the identified non-coding variants. Together, the combination of rarity, predicted coding impact, evolutionary conservation, strong gene-level constraint and prior memory-related biology supported *MYCBP2* as the highest-priority candidate for functional investigation. This prioritization does not exclude contributions from the other candidates, particularly given the unknown genetic architecture and penetrance of HSAM.

Sanger sequencing confirmed the *MYCBP2* variant in JN and its absence in both parents. The variant is a single nucleotide substitution, NC_000013.11:g.77251230C>A (GRCh38), corresponding to NM_015057.5:c.2302G>T and predicting p.(Val768Phe) in MYCBP2 (Fig. 1d,e).

*MYCBP2* is strongly constrained against both predicted loss-of-function and missense variation in gnomAD v2.1.1 (pLI = 1; pLoF o/e = 0.08 [0.05–0.12]; missense Z = 6.05; missense o/e = 0.65 [0.63–0.68]), whereas synonymous variation is not depleted (Z = 0.01; o/e = 1.00 [0.94–1.06]). *MYCBP2* is involved in axon guidance and synapse formation and is highly conserved across species^20, 23, 24^. The identified variant affects the valine residue at codon 768 of the human reference sequence in the RCC1-like domain (RLD) located in the N-terminal part of the protein. This residue is conserved across vertebrates (e.g., non-human primates, *Mus musculus, Canis familiaris, Xenopus tropicalis, Danio rerio*) and invertebrates (e.g., *Drosophila melanogaster, Caenorhabditis elegans*) (Fig. 1e). A subsequent check of the updated gnomAD v4 database confirmed the absence of the MYCBP2 variant in >700,000 exomes.

To complement the initial trio-WES analysis, we performed long-read whole-genome sequencing of JN as an exploratory search for structural-variant calls with potential direct gene-level consequences. This analysis was not designed to establish definitive inheritance and does not rule out a contribution from structural variation (see Methods). Among 23,407 SV calls ≥50 bp, 29 non-breakend (BND) calls overlapped annotated exonic regions and showed less than 50% reciprocal overlap with SVs in the selected population-reference callsets. BND calls were assessed separately for exonic or splice-region overlap, adding one candidate BND call. The resulting 30 candidate calls were evaluated using trio-WES alignment patterns and, for deletion and duplication calls, GATK gCNV calls and normalized copy-ratio profiles. No call showed sufficiently reliable proband-specific evidence to meet Category 1 (defined as proband-specific evidence concordant with the long-read call and absent in both parents despite informative coverage); 10 were classified as unresolved or not corroborated by proband-specific WES evidence, and 20 showed evidence suggestive of a shared event with at least one parent (Supplementary Table 4; Supplementary Data 1). Accordingly, the long-read analysis did not identify an SV call that warranted prioritization over *MYCBP2* V768F for functional follow-up.

### Effect of CRISPR-edited MYCBP2/rpm-1 variant on long-term memory in C. elegans

To test whether the identified *de novo MYCBP2* variant causally affects long-term memory, we first introduced the variant in *C. elegans* and assessed long-term memory. *C. elegans* harbours a single highly conserved orthologue of *MYCBP2*, known as *rpm-1*, which plays a crucial role in regulating axonal development and synaptic strength^20^. Using CRISPR/Cas9 genome editing, we introduced the variant into the corresponding conserved residue of RPM-1, generated *rpm-1(V682F)* animals, and confirmed the presence of the edit by sequencing. The missense variant is located in the N-terminal RCC1-like domain (RLD) of *rpm-1* (Fig. 2a). Because missense variants in *C. elegans* can exhibit temperature-dependent phenotypic effects^29–31^, we conducted experiments under two different temperature conditions.

**Fig. 2.**
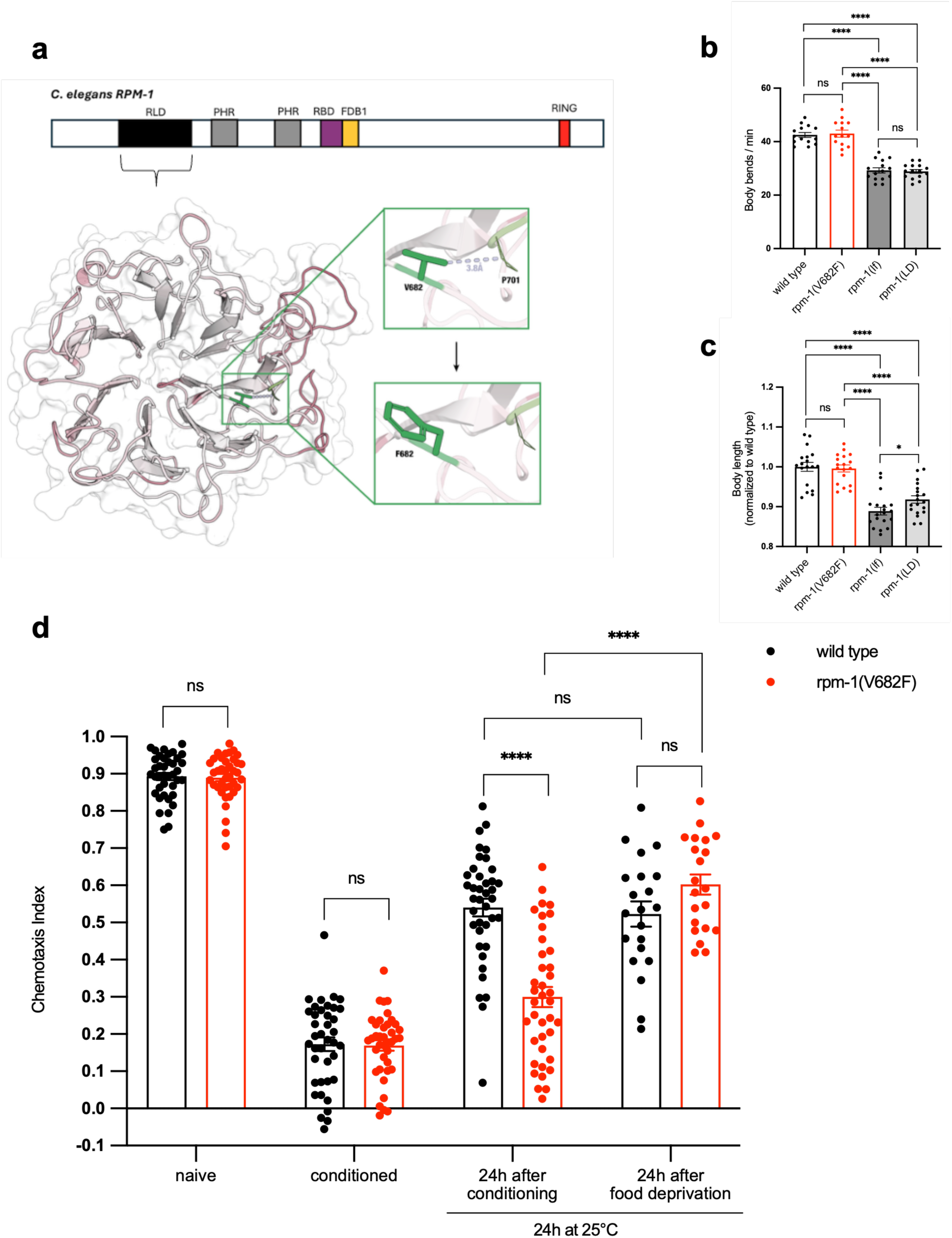
Impact of the mutated MYCBP2/RPM-1 residue on developmental and behavioral measurements in *C. elegans*. **a**, Schematic representation and domain structure of *C. elegans* RPM-1. Template PDB ID 8gqe was used for RPM-1 RLD domain structural modeling. Structure quality is depicted on a red to white scale using QMEAN scoring. Residue V682 and the mutated version F682 are coloured green and depicted as sticks. A predicted hydrophobic interaction of V682 with P701 (green lines) is shown as a light blue dashed line. **b**,**c**, Locomotor behavior, i.e. body bends/min (**b**), and body length (**c**) of the adult N2 WT strain and three adult *rpm-1* mutant strains (*rpm-1(V682F)*, loss-of-function *rpm-1(lf)* (i.e., *rpm-1(utr93[3xstop+frameshift])*, see Methods), and an RPM-1 ligase-dead (*rpm-1(LD)*) site-directed mutant strain carrying variants within the RING domain (C3535A, H3537A, and H3540A) that impair catalytic activity^32^). Dots represent single worms. **d**, Chemotaxis indices towards DA of WT and *rpm-1(V682F)* worms under four conditions: naïve (unconditioned), conditioned (immediately after 1h of food deprivation in the presence of DA), 24 h after conditioning, 24 h after 1h of food deprivation in the absence of DA. For the last two conditions, worms were kept at 25°C during 24 h. Dots represent single plates. For all bar plots in **b-d**, bars and error bars represent means and s.e.m., respectively. All *P* values in **b-d** are two-sided (non-parametric Wilcoxon-Mann-Whitney test). ns: *P* ≥ 0.05, \**P* < 0.05, \*\*\*\**P* < 0.0001.

Animals were tested in an olfactory learning and memory assay, in which worms are first trained to avoid diacetyl (DA), which they are normally attracted to since it signifies food, by pairing it with an aversive condition of food deprivation. Because olfactory conditioning relies on normal detection of volatile attractants, we first tested the chemotaxis of *rpm-1(V682F)* animals toward different odorants. The chemotaxis of *rpm-1(V682F)* mutants to two different volatile attractants (DA, isoamyl alcohol (IAA)) and a repellent (octanol) was comparable to the response of the wild-type (WT) N2 strain, regardless of the growth temperature (Supplementary Fig. 2a). Furthermore, both WT and *rpm-1(V682F)* mutants had normal locomotor behaviour (Fig. 2b) and responded similarly to food (Supplementary Fig. 2b), indicating that *rpm-1(V682F)* mutants have no obvious motor or sensory defects. In addition, WT animals and *rpm-1(V682F)* mutants exhibited similar body lengths (Fig. 2c), suggesting that the variant does not affect normal growth of these worms. In contrast, *rpm-1* loss-of-function mutants (*rpm-1(lf)*) and animals harbouring variants that severely affect RPM-1 ubiquitin ligase activity (*rpm-1(LD)*, i.e., ligase-dead)^32^ showed decreased locomotor activity and reduced body length (Fig. 2b,c). The locomotor impairments of *rpm-1(lf)* and *rpm-1(LD)* precluded interpretable assessment in the chemotaxis-based learning and memory assay, which requires animals to navigate towards or away from an odor source. Memory experiments therefore compared WT and *rpm-1(V682F)* animals, which showed comparable locomotor and baseline chemosensory responses.

In the olfactory learning assay, both unconditioned WT and *rpm-1(V682F)* mutant animals demonstrated robust chemotaxis towards DA (Fig. 2d). Moreover, following a 1-hour period of food deprivation in the presence of DA (aversive conditioning), both WT and *rpm-1(V682F)* mutants exhibited normal associative learning (Fig. 2d). In a control experiment, both strains exhibited similar attraction to DA after food deprivation and after exposure to DA alone (in the presence of ample food) (Supplementary Fig. 2b).

To assess the impact of the *V682F* variant on long-term associative memory (LTAM), we evaluated animals’ retention of conditioned behaviour over time as described previously^33^. We observed significant differences in LTAM retention between *rpm-1(V682F)* mutant animals and WT worms following a 24-hour delay period at 25°C (*P* = 8×10^-8^; Fig. 2d). After 24 hours, WT animals demonstrated a complete loss of LTAM retention, as their chemotaxis towards DA was almost identical to that of unconditioned animals of the same age (Fig. 2d, last condition, i.e., 24 h after food deprivation for 1h in the absence of DA). In contrast, the chemotaxis towards DA of *rpm-1(V682F)* mutants 24 h after conditioning was significantly lower than that of unconditioned *rpm-1(V682F)* mutants of the same age (*P* = 2×10^-7^; Fig. 2d, last condition, i.e., 24 h after food deprivation for 1h in the absence of DA). No difference in LTAM between conditioned strains was observed following a 24-hour delay period at 20°C (Supplementary Fig. 2c). Together, the V682F variant enhances long-term memory retention in *C. elegans* in a temperature-dependent manner.

### Axonal development in CRISPR-edited MYCBP2/rpm-1 C. elegans mutants

Because MYCBP2/RPM-1 has established roles in axon development^20^, we next examined whether the memory-enhancing V682F variant also affects axon termination in *C. elegans.* In nematodes, disruption of RPM-1 function leads to axon termination defects, characterized by the overgrowth of sensory and motor axons^34, 35^. This phenotype is likely attributable to altered ubiquitination at axon termination sites^26, 32^. In *C. elegans*, C-terminal mutations compromise RPM-1 ubiquitin ligase activity and result in axonal abnormalities^26^.

We tested the effect of JN’s N-terminal variant on axon termination in the anterior lateral mechanosensory (ALM) and posterior lateral mechanosensory (PLM) neurons utilizing a strain that expresses GFP in these neurons (muIs32 [mec-7p::GFP])^36^. Conventional *rpm-1* loss-of-function mutants (*rpm-1(lf)*) and animals harbouring variants that severely affect RPM-1 ubiquitin ligase activity (*rpm-1(LD)*) displayed severe axon termination defects (Supplementary Fig. 2d). In contrast, axon termination in *rpm-1(V682F)* mutants was not impaired under physiological growing conditions (Supplementary Fig. 2d) and was comparable to that of WT animals under conditions known to affect axon development (Supplementary Fig. 2d). These results indicate that the V682F variant does not alter *in vivo* axon development in this *C. elegans* model.

### Long-term memory in heterozygous Mycbp2 knock-in mice

Having shown that the *MYCBP2* variant enhances long-term memory in an invertebrate model organism, we next asked whether this variant enhances long-term memory in mice. We therefore generated a CRISPR knock-in mouse carrying the orthologous *MYCBP2* mutation (see Methods and Supplementary Data 2). Using two-cell embryo microinjection and homology-directed repair, we introduced a valine-to-phenylalanine substitution at the V768 position in the mouse *Mycbp2* gene (the murine equivalent of human V768F, referred to here as *Mycbp2*^V>F). Mice heterozygous for the variant (*Mycbp2*^+/V>F), matching JN’s heterozygous state, were viable and showed no abnormalities or neurological deficits in the SHIRPA primary observation screen (See Methods and Supplementary Data 3).

First, we tested spatial memory in the Barnes maze (Fig. 3a). Mice (heterozygous N = 19 (9 female), mean age (days) at start of training ± s.d.: 99 ± 8; wild-type N = 20 (10 female), mean age (days) at start of training ± s.d.: 102 ± 10) were trained to navigate to one of 20 holes arranged around the perimeter of a circular platform, leading to an escape nest, using visual cues. Time spent in the target zone area during subsequent probe tests was quantified as an indicator of spatial memory performance. Training was conducted over 4 days, followed by probe tests 1 and 8 days following the final training session. Memory for the target location did not decline significantly across these retention intervals (main effect of retention interval: repeated measures ANOVA *P* = 0.11). Performance was comparable between genotype groups, with no main effect of genotype (*P* = 0.57) and no genotype × retention interval interaction (*P* = 0.8) (Fig. 3a).

**Fig. 3.**
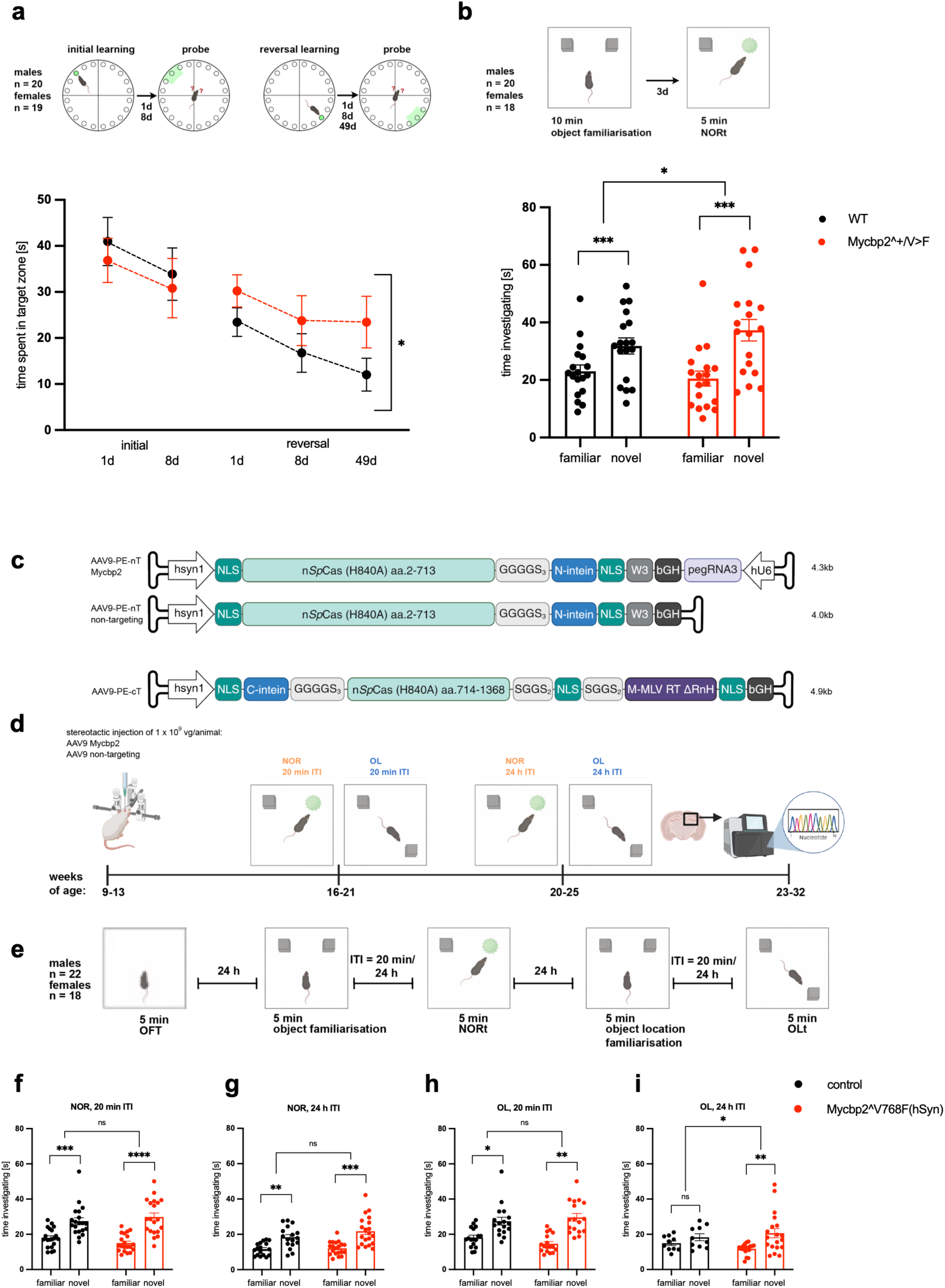
Long-term memory in heterozygous Mycbp2^+/V>F knock-in mice and after hippocampal prime editing to introduce MYCBP2^V768F. **a**, Barnes maze design for initial and reversal learning, with probe tests at 1 and 8 days after initial training and at 1, 8, and 49 days after reversal training (*n* as indicated). Bottom, time spent in the target zone (initial) or new target zone (reversal) during probe tests for wild-type (WT; black) and Mycbp2^+/V>F mice (red). Dots and error bars represent means and s.e.m, respectively. The asterisk indicates a significant effect of genotype across reversal probe tests. **b,** Novel object recognition (NOR) paradigm with a 3-day retention interval (top, *n* as indicated) and quantification of time investigating familiar versus novel objects during the test phase (bottom) for WT (black) and Mycbp2^+/V>F mice (red). For the statistical comparison of genotype groups, the discrimination index (DI) served as dependent variable. Mice with total exploration time (TET) < 20 sec (*n* = 2) were excluded from the analysis (see Methods). **c,** Schematic representation of the AAV9 vectors encoding the N-and C-terminal fragments of the intein-split prime editor under the human synapsin 1 (hSyn1) promoter. **d,** Timeline of the *in vivo* prime editing experiments, showing bilateral AAV9 delivery to the dorsal hippocampus, subsequent NOR and object location (OL) testing with 20-min and 24-h inter-trial intervals (ITI), and brain collection for quantification of on-target editing. Ages are indicated in weeks. Created in BioRender (https://BioRender.com/). **e,** Behavioral schedule for the gene-editing cohort, including open field testing and subsequent NOR and OL tasks (20 min and 24 h ITI), with n as indicated. **f,g,** NOR performance after a 20-min ITI (**f**) and 24-h ITI (**g**) shown as time investigating familiar versus novel objects for control mice (black) and hippocampus-edited Mycbp2^V768F(hSyn) mice (red). For statistical comparisons of genotype groups (**f**), the discrimination index (DI) served as dependent variable (see Methods). For statistical comparisons of genotype groups (**g**), the raw exploration difference score served as dependent variable, with TET as a covariate. Mice with total exploration time < 20 sec (*n* = 2) were excluded from the analysis (see Methods). **h,i,** OL performance after a 20-min ITI (**h**) and 24-h ITI (**i**) shown as time investigating the object in the familiar versus displaced (novel) location for control (black) and hippocampus-edited Mycbp2^V768F(hSyn) mice (red). For statistical comparisons (**h**), the raw exploration difference score served as dependent variable, with TET as a covariate. Mice with total exploration time < 20 sec (*n* = 6) were excluded from the analysis (see Methods). For statistical comparisons of genotype groups (**i**), the raw exploration difference score served as dependent variable, with TET as a covariate. Mice with TET < 20 sec (*n* = 11) were excluded from the analysis (see Methods). Furthermore, one animal was excluded based on the predefined outlier criterion for TET (see Methods). Data (**b, f-i**) are shown as scatter plots (mean with s.e.m.) with individual animals overlaid as dots. Statistical significance is indicated in the panels (\**P* < 0.05, \*\**P* < 0.01, \*\*\**P* < 0.001, \*\*\*\**P* < 0.0001; ns, not significant); no significant genotype x sex interactions were observed in any model (all *P* > 0.05); all tests were two-sided and details are provided in Methods.

Whereas no genotype effect was detected after the initial learning phase, a difference emerged after reversal learning, a more challenging condition, in which mice had to learn a new escape location under shorter training and with extended retention intervals. In this phase, the escape hole was relocated to the opposite quadrant, and mice underwent 3 days of reversal training to acquire the new location. Reversal probe tests were performed 1 and 8 days after the final reversal training session, mirroring the time points used after initial acquisition. An additional probe test was conducted 49 days after reversal training, representing an extended retention interval. In the reversal phase, memory performance declined across retention intervals (main effect of retention interval: ART ANOVA *P* = 0.0003). *Mycbp2^+/V>F* mice spent more time in the target zone than wild-type littermates (main effect of genotype: *η*^2^ = 0.15, *P* = 0.024; nparLD sensitivity analysis: mATS *P* = 0.031), with no genotype × time interaction (*P* = 0.7), indicating a consistent genotype-related performance difference across retention intervals (Fig. 3a).

In the same cohort, we next assessed non-spatial recognition memory using the novel object recognition (NOR) test (Fig. 3b). Mice (heterozygous N = 19 (9 female), mean age (days) at start of testing ± s.d.: 172 ± 7; wild-type N = 19 (9 female), mean age (days) at start of testing ± s.d.: 175 ± 8) explored two identical objects in an open-field arena for 10 min during the training phase and were then returned to their home cages. Following a 3-day retention interval the animals were reintroduced to the arena containing one familiar object and one novel object, and object exploration time was quantified. Both *Mycbp2^+/V>F* mice and wild-type littermates showed intact memory at this retention interval, spending significantly more time investigating the novel than the familiar object (paired-samples t-test *P* = 0.0005; *P* = 0.0006, respectively, Fig. 3b). However, *Mycbp2*^+/V>F mice exhibited a greater preference for the novel object compared to wild-type littermates (ANOVA, *η*^2^ = 0.12, *P* = 0.047, Fig. 3b). Together with the Barnes maze findings, these results indicate that heterozygous *Mycbp2* knock-in mice exhibit enhanced long-term memory performance across spatial and non-spatial memory tasks.

#### Memory effect of hippocampus-specific prime editing of the MYCBP2 variant in adult mice

Because the long-term memory phenotype in the germline knock-in mouse model could arise from developmental mechanisms as well as from direct effects of the *MYCBP2* variant in the adult brain, we introduced the variant postnatally via prime editing in dorsal hippocampal neurons of adult mice (see Methods; Fig. 3c,d; Supplementary Fig. 3, 4). This allowed us to test whether adult hippocampal expression of the variant, specifically in neurons, is sufficient to enhance long-term memory. To this end, we employed behavioural tasks with differing degrees of hippocampal dependence (Fig. 3d,e). Specifically, we used the NOR test alongside the object location (OL) test, which relies more strongly on hippocampal processing^37–39^.

In the NOR test, mice (hippocampus-edited Mycbp2^V768F(hSyn) N = 20 (9 female), mean age (days) at start of the experiment ± s.d.: 127 ± 12; control N = 20 (9 female), mean age (days) at start of the experiment ± s.d.: 126 ± 13) explored two identical objects in an open-field arena for 5 min and were then returned to their home cages. Memory was assessed after either a short (20 min) or long (24 h) inter-trial interval (ITI) by replacing one object with a novel one and quantifying preferential exploration of the novel object (Fig. 3d,e). Both Mycbp2^V768F(hSyn) and control mice injected stereotaxically with a non-targeting pegRNA (Fig. 3c,d) exhibited intact memory at the 20 min ITI (paired-samples t-test *P* = 7×10^-6^; P = 0.001, respectively, Fig. 3f) and at 24 h ITI (paired-samples t-test *P* = 0.0003; *P* = 0.003, respectively, Fig. 3g), with no significant difference between genotype groups at either delay (ANOVA *P* = 0.093; *P* = 0.9, respectively, Fig. 3f,g).

In the OL test, mice (hippocampus-edited Mycbp2^V768F(hSyn) N = 20 (9 female), mean age (days) at start of the experiment ± s.d.: 128 ± 12; control N = 20 (9 female), mean age (days) at start of the experiment ± s.d.: 127 ± 13) explored two identical objects in an open-field arena during the object familiarisation phase. Memory for the spatial configuration was assessed after either a short (20 min) or long (24 h) ITI by relocating one object and quantifying preferential exploration of the displaced object (Fig. 3d,e). Mycbp2^V768F(hSyn) mice and control littermates exhibited intact OL memory at the 20 min ITI, spending significantly more time investigating the object in the displaced location than in the familiar location (paired-samples t-test *P* = 0.002; *P* = 0.025, respectively, Fig. 3h), with no significant difference between groups (ANCOVA *P* = 0.13, Fig. 3h). At the 24 h ITI, control mice no longer exhibited preference for the displaced object (paired-samples t-test *P* = 0.2, Fig. 3i). In contrast, Mycbp2^V768F(hSyn) mice exhibited intact OL memory at the 24 h ITI (Wilcoxon *P* = 0.009, Fig. 3i). The difference between genotype groups was significant (ANCOVA, *η*^2^ = 0.19, *P* = 0.030, Fig. 3i). In this analysis, one animal was excluded based on the predefined outlier criterion for total exploration time (see Methods); the genotype effect remained significant when this animal was included (ANCOVA *P* = 0.034).

These results demonstrate that introducing the human *MYCBP2* variant into the adult dorsal hippocampus is sufficient to enhance hippocampus-dependent long-term spatial memory in the OL test, without affecting short-delay performance. By contrast, the same hippocampal editing did not alter object recognition performance at either short or long delays under our NOR test conditions.

## Discussion

JN is an individual with previously reported HSAM, whose exceptional autobiographical memory was confirmed by his near-perfect performance on the 10 Dates Quiz (29 out of 30), while his scores on standard cognitive tests across multiple domains were within the normal range (Fig. 1a). This profile indicates that his mnemonic superiority is selective to autobiographical recall and is not accompanied by broader cognitive alterations. Subsequent whole-brain volumetric analysis revealed exceptionally large volumes (Z > 3) specifically in multiple hippocampal subregions known to support autobiographical memory (Fig. 1b,c). Although the precise localization of the structural changes should be interpreted cautiously given the limitations of current MRI techniques^40^ and automatic segmentation algorithms, the outlying volumes were confined to the right parasubiculum, right head of the granule cell and molecular layers of the dentate gyrus, right head of the CA4 region, and right head of the CA3 region. No other cortical or subcortical regions exhibited volumes exceeding Z = 3. Of note, among the 7755 UK Biobank participants, none demonstrated extreme volumes in all four sub-regions of the hippocampal formation, a feature present in JN. Prior studies in individuals without HSAM have found that larger volumes in hippocampal subregions correlate with better autobiographical memory performance^15–19^. For example, bilateral CA2/3 volume is positively associated with autobiographical recall ability^16^, and volumes of the dentate gyrus, CA2/3, and subiculum correlate with autobiographical episodic memory performance in young adults^18^. Further, the amount of autobiographical detail recalled has been linked to the volume of the pre/parasubiculum and CA3/2 regions^15^. Thus, JN’s hippocampal anatomy aligns with his extraordinary memory capacity and echoes structure-function relationships observed in the broader population. Consistent with this, functional MRI studies of other HSAM individuals have reported enhanced hippocampal– prefrontal connectivity during autobiographical recall^3^, underscoring the central role of the hippocampus in this superior memory phenotype.

We found JN to carry a rare *de novo* missense variant in *MYCBP2* (V768F) that is absent from >700,000 sequenced exomes. To test its causal impact on memory, we introduced the homologous variant into model organisms. In *C. elegans*, animals carrying the V>F variant showed a temperature-dependent enhancement of long-term memory. In mice, independent experiments demonstrated that the variant enhances long-term memory retention. Specifically, heterozygous knock-in mice carrying the variant showed significantly improved object recognition memory following a 3-day retention interval and significantly improved memory for the reverse Barnes maze escape location across probe tests extending to 49 days after training compared with wild-type littermates. Second, adult mice with the variant introduced postnatally via prime editing in dorsal hippocampal neurons also displayed better long-term memory: these mice retained, unlike control mice, object location memory after a 24-hour delay, while both groups performed equivalently at a 20-minute delay. By contrast, the same manipulation did not alter NOR performance at either short or long delays. This dissociation is consistent with the recognition-memory literature showing that standard NOR is often preserved after hippocampal lesions, whereas memory for object location is more strongly dependent on hippocampal, particularly dorsal hippocampal, circuitry. At the same time, hippocampal contributions to object recognition can emerge under specific task conditions, so our data are best interpreted as showing task-selective enhancement under the present NOR and OL conditions rather than a complete absence of hippocampal involvement in NOR^37–39^. Importantly, our NOR protocol used the standard arrangement of two identical objects during the sample phase and preserved object locations at test, a design that has been argued to minimize additional spatial or relational demands and therefore to reveal more clearly the relative sparing of standard object recognition after hippocampal disruption^37–39^. An alternative explanation for the dissociation between the OL and NOR findings is that the behavioural effect of the variant emerges preferentially under conditions in which memory is weaker or closer to the forgetting threshold. In the present experiments, this appears to have been the case for OL at the 24-h delay, whereas NOR memory remained detectable in control mice at the same interval. Accordingly, the dissociation between tasks may reflect differences in baseline memory strength and assay sensitivity, in addition to possible differences in the underlying neural circuitry. Together, the mouse experiments indicate that the variant produced selective enhancements in long-delay memory performance, particularly under conditions with higher retention demands. The concordant memory improvements observed in nematode and mouse support a conserved role for the *MYCBP2* V768F variant in enhancing memory. The demonstration of better long-term memory in two mouse models executed in different laboratories further supports the robustness of the variant’s impact.

*MYCBP2* encodes a large, evolutionarily conserved protein (also known as Phr1 in mammals, RPM-1 in nematodes, and Highwire in flies) that functions as an E3 ubiquitin ligase and a multifaceted signalling hub. It contains several recognizable domains (Fig. 1e) and has critical roles in axon development and synaptic function^20, 23–25^. In humans, eight individuals carrying distinct *de novo MYCBP2* variants have been reported with a neurodevelopmental phenotype including developmental delay/intellectual disability, autistic features, epilepsy and corpus callosum abnormalities; functional analyses supported partial or strong loss-of-function effects of the tested alleles^26^. V768F lies in the N-terminal RCC1-like domain, whereas the missense variants reported by AlAbdi et al. ^26^ affect more C-terminal residues.

In our assays, *rpm-1(V682F)* did not reproduce the locomotor, growth or axon-termination defects observed in the strong loss-of-function and ligase-dead strains. Moreover, introducing V768F into dorsal hippocampal neurons of adult mice enhanced long-delay memory, demonstrating that expression during early development is not required for this effect. These observations are compatible with a variant-specific effect distinct from the previously described MYCBP2 loss-of-function spectrum.

Complete loss of MYCBP2 (or its homologues) in model organisms leads to severe neurodevelopmental abnormalities: rpm-1 null *C. elegans* have aberrant neuronal morphology and locomotor defects^23, 34, 41^, *Drosophila highwire* mutants show excessive axon branching and motor dysfunction^24^, and mice lacking Mycbp2/Phr1 die perinatally with major axonal pathologies^42, 43^. Interestingly, partial perturbations of this pathway can enhance memory capacity and neuroprotection in animal models: *Drosophila highwire* loss-of-function mutants exhibit facilitated long-term memory formation^44^, and conditional *Mycbp2* deletion in adult mice leads to prolonged survival of injured axons in both central and peripheral nervous systems^45^. These findings highlight the complex, dose-dependent role of PHR proteins in neural circuit stability and plasticity, raising the possibility that a specific hypomorphic alteration of human MYCBP2 could enhance memory retention while avoiding detrimental developmental effects. However, the absence of strong loss-of-function phenotypes does not establish the molecular direction of the V768F effect. Whether this variant causes partial loss of function, gain of function or a qualitative change in protein interactions or substrate specificity remains an object of future research. Furthermore, we observed that the memory-enhancing effect of the *MYCBP2* variant in *C. elegans* was temperature-dependent, indicating that its phenotypic expression is modulated by environmental conditions in this model.

Our study has the following limitations: The human genetic evidence is based on a single individual. Although introducing the homologous *MYCBP2* variant enhances memory in *C. elegans* and mice, these experiments do not establish that V768F caused JN’s HSAM or that HSAM is a monogenic *MYCBP2*-related trait. We therefore consider V768F a biologically compelling candidate contributor to the human phenotype. Evidence from additional unrelated individuals or informative families carrying *MYCBP2* variants in association with comparable phenotypes would strengthen the proposed human gene-phenotype relationship. Why JN appears to show a selective enhancement of autobiographical memory and whether this pattern depends on his genetic background or human-specific cognitive and neural factors remain open questions. The precise molecular and circuit mechanisms by which this missense change improves memory in model systems also remain unknown. The affected amino acid lies in a poorly characterized N-terminal region of the protein, and we speculate that V768F may subtly alter MYCBP2’s interactions or substrate specificity to promote synaptic stability (for example, by slowing the turnover of memory-related proteins). Such a mechanism has precedent in memory biology. For example, KIBRA/WWC1 stabilizes the memory-related kinase PKMζ by protecting it from proteasomal degradation, while WWC1/2 similarly stabilize AMOT family proteins through deubiquitination; persistent KIBRA-PKMζ interactions can maintain long-term potentiation and memory despite ongoing molecular turnover^46–49^. Conversely, retrieval of established memories engages ubiquitin-proteasome-dependent degradation of synaptic proteins, a process required for memory destabilization and subsequent reconsolidation or updating in hippocampal and amygdala-dependent memory paradigms^50–54^. These findings illustrate how regulated synaptic protein turnover can influence memory persistence and lability, although whether MYCBP2 V768F acts through such a mechanism remains unknown. Further work will be required to elucidate how this single amino acid change enhances memory. Finally, it remains to be seen whether the variant’s effect in model organisms extends to other forms of memory or cognitive processes beyond those tested, and whether chronically enhanced memory retention carries any trade-offs for neural function. Addressing these questions will be important for fully understanding MYCBP2’s role in memory.

In conclusion, our study identifies a rare de novo *MYCBP2* variant in an individual with HSAM and demonstrates that the homologous substitution enhances long-term memory in *C. elegans* and mice. These findings lay the groundwork for mechanistic dissection of MYCBP2’s involvement in memory, and they raise the possibility that insights from such rare, memory-enhancing variants could inspire new strategies for combating memory loss.

## Data Availability

All data produced in the present study are available upon reasonable request to the authors.

## Acknowledgements

We are deeply grateful to JN and his family; without their generous contribution and dedication to research, this work would not have been possible. This research has been conducted using the UK Biobank Resource under Application Number 101531. Funding to A.P. and D.J.-F.d.Q. was provided by the University of Basel and the Novartis Forschungsstiftung (FreeNovation Grant nr. FN19-28). Funding to S.M. and P.B.-S. was provided by the German Federal Ministry of Research, Technology and Space (BMFTR, 01EJ2502A and ERA-NET NEURON 01EW2305), and the German Research Foundation (DFG, Project-ID 424778381-TRR 295 ReTune and EXC-2049-390688087 NeuroCure) and supported by Charité 3^R^ – Replace | Reduce | Refine.

## Author contributions

Conceptualization: A.P. and D.J.-F.d.Q. Methodology, formal analysis and investigation: A.P., D.N., D.B., J.P., A.A., A.K.R.L.P., P.M., M.N., V.F., V.G., N.S., V.V., D.C., A.B., A.S., N.B., C.R., M.L., D.Bo., J.D., T.S., O.B., K.H., P.P., B.B., V.T., C.E.L.S., J.L.Mc.G., G.S., J.B., C.D. and D.J.-F.d.Q. Resources: A.P., O.B., M.L., S.M., P.B.-S., P.P., V.T., C.E.L.S., J.L.Mc.G., G.S., J.B., C.D. and D.J.-F.d.Q. Data curation: D.N., D.B., J.P., A.A., V.F., N.S., V.V., D.C., M.L. and D.Bo. Writing—original draft: A.P., D.N., D.B., J.P., A.A., V.F., N.S., D.C., M.L., D.Bo., V.V., E.M.C.S., P.P., C.E.L.S., J.L.Mc.G., G.S., J.B. and D.J.-F.d.Q. Writing—review, revision, and editing: A.P., D.N., D.B., J.P., A.A., A.K.R.L.P., P.M., M.N., V.F., N.S., D.C., A.B., C.R., M.L., D.Bo., V.V., A.S., N.B., V.G., S.M., N.G., J.D., T.S., O.B., J.G., E.M.C.S., K.H., B.B., S.C., P.B.-S., P.P., V.T., C.E.L.S., J.L.Mc.G., G.S., J.B., C.D. and D.J.-F.d.Q. Supervision: A.P. and D.J.-F.d.Q.

## Competing interests

The authors declare no competing interests.

## Materials & Correspondence

Correspondence and material requests should be addressed to A.P., D.J.-F.d.Q., C.D., J.B., or G.S.

## Methods

### Brain imaging

#### UK Biobank participants

Data for the control population were obtained from the UK Biobank through research application number 101531. Informed consent was obtained by the UK Biobank for all participants^55^. The data release included 502,413 participants. To match the age and sex of our HSAM participant, we restricted the sample to participants fulfilling the following predefined criteria:

- participated in the baseline visit as well as in the imaging visit (instance 2)
- self-reported overall health rating “Excellent”, “Good” or “Fair” (data-field 2178)
- sex “Male” (data-field 31)
- age at imaging visit greater than 58 and lower than 68 years old (data-field 21003) This corresponded to 9,356 participants, of whom 7,838 had complete imaging data.

#### HSAM participant: MRI acquisition and pre-processing

Structural MRI data was acquired on a 3T Siemens Magnetom Prisma scanner with a 20-channel RF receive head-neck coil, following the UK Biobank imaging protocol^56^. Recent evidence confirms high inter-site reliability of the Skyra-based UK Biobank imaging protocols on the Prisma platform, particularly for grey matter phenotypes such as volume and cortical thickness, which are central to this study. Protocols were adapted to minimize variability between Skyra and Prisma scanner data^57^.

The T1 structural image was a 1 mm isotropic resolution three-dimensional magnetization-prepared rapid-acquisition gradient-echo (3-D MPRAGE) acquisition with the following parameters: field of view = 256×256 mm; time of acquisition = 4 min 53 s; repetition time = 2000 ms; inversion time = 880 ms; echo time = 2.03 ms; flip angle = 8°; 208 sagittal slices. The T2-weighted image consisted of a fluid-attenuated inversion recovery (FLAIR) image, with the 3D SPACE optimized readout and the following parameters: field of view = 256×256 mm; time of acquisition = 5 min 52 s; repetition time = 5000 ms; inversion time = 1800 ms; echo time = 397 ms; flip angle = 120°; slice thickness = 1.05 mm; 192 sagittal slices.

Data were pre-processed using the pipeline for brain imaging processing of the UK Biobank^58^ (https://git.fmrib.ox.ac.uk/falmagro/uk_biobank_pipeline_v_1.5), via an adapted Docker container (https://git.fmrib.ox.ac.uk/paulmc/fbp-dockerfile, https://github.com/dcoynel/fbp-dockerfile). Correction of gradient distortion was performed as a first step of the T1 pipeline, using the scanner’s own gradient coefficients (*coeff.grad*)^58^.

#### Image-derived phenotypes

Of interest for this project were the image-derived phenotypes (IDPs) provided by the cortical and subcortical pipeline from FreeSurfer v6.0.0^11–13^. Both the T1 and T2 images were used as input for FreeSurfer^14^. The resulting IDPs consisted of cortical and subcortical volumes, cortical thickness and cortical surface area. Concerning the cortical data, we included data from all cortical parcellations available: the Desikan atlas, the Desikan-Killiany–Tourville atlas (denoted DKT), the pial surface parcellation Desikan-Killiany atlas (denoted pial) and the Destrieux atlas (denoted a2009s). The following subcortical structures were further sub-divided: amygdala (nuclei volumes), hippocampus (subfield volumes), thalamus (nuclei volumes), brainstem (substructure volumes). Subjects without estimated total intracranial volume were excluded (data-field 26521; N=22). The following IDPs were excluded from further analyses because more than 5% of the UK Biobank participants had a value of 0: volume of 5th-Ventricle (data-field 26525) and volume of non-WM-hypointensities (data-field 26529). For a given IDP, a subject was excluded if he had a value that was outside the population’s mean +/-10 standard deviations. This was the case 138 times, distributed over 61 subjects and 70 IDPs.

The final data used for statistical analyses consisted of 1070 IDPs for 7755 UK Biobank subjects.

#### MRI statistical analyses

The data were further corrected for total brain volume and z-transformed. For correcting for total brain volume, we considered for each IDP the residuals of a linear model including only the intercept and the total intracranial volume (TIV). Those values were then Z-transformed using the mean and standard deviation of the UK Biobank participants only. The resulting Z-scores then represent the amount of standard deviation a given subject is from the TIV-corrected control population’s mean. The analysis focused on identifying whether any of JN’s brain IDPs exceeded Z = 3, indicating metrics that can be empirically considered as extremely large values^10^ (i.e., lying in the outer 0.13% of the respective population distribution). Supplementary Table 2 reports all Z values (positive and negative).

### Human genetics methods

#### Whole exome sequencing (WES)

WES for JN and his parents was performed together with that of 253 unrelated control individuals (healthy young adults of European ancestry). Participants in the internal WES cohorts were not formally assessed for HSAM. Blood sampling, DNA isolation and library preparation was done as described previously ^59^. WES was done on the Illumina HiSeq platform (paired-end reads). Samples were processed in a double-randomized manner. Samples were randomly pooled and each pool was sequenced in multiple lanes. For each sample, over 9 Gb of sequence were generated. The SureSelect XT Human All Exon V6 + UTR kit (Agilent) was used for targeting regions. Each sample’s sequence was mapped to the hg19 human reference genome, using BWA (Burrow-Wheeler Alignment) v0.7.12 ^60^. Duplicates were flagged with Picard v1.135. Local realignment and base quality score recalibration were performed with the Genome Analysis Toolkit (GATK) ^61^ v3.4-0, following the standard GATK protocol (https://gatk.broadinstitute.org). Haplotype Caller was used for variant calling and recalibration. We chose 99% sensitivity for a variant to be ‘true’ based on an adaptive error model and filtered out false-positive variants using this threshold. Variants with any missing values were also filtered out.

#### De novo variant identification

*De novo* variant identification based on WES data is highly susceptible to false positive findings. Therefore, we took the following steps to nominate potential *de novo* variants with high confidence. First, we ran the Genotype Refinement Pipeline (https://gatk.broadinstitute.org/hc/en-us/articles/360035531432-Genotype-Refinement-workflow-for-germline-short-variants), GATK 3.4-0, in order to improve the accuracy of genotype calls and to filter calls that are not reliable enough for downstream analysis. This workflow is recommended as particularly useful for analysis concerning transmission or *de novo* origin of variants in a family. Specifically, the “CalculateGenotypePosteriors” tool was used to calculate the posterior probabilities for each sample at each variant site, including family priors, i.e., pedigree data, for the HSAM trio. Next, the “VariantFiltration” tool was used to identify genotypes with GQ >= 20 based on the posteriors, indicating that there is a 99% chance that the genotype in question is correct. Next, “VariantAnnotator” was used to tag possible *de novo* variants. High confidence *de novo* sites were defined as having GQs >= 20.

To further refine the *de novo* variants search, we also applied another *de novo* variants filtering pipeline, independently from the GATK pipeline. We used SAMtools 1.9 for variant calling and VarScan v2.4.1 ^62^ for identification of *de novo* variants. For comparison of the two pipelines, see refs. ^63, 64^. Variants identified as high-confidence *de novo* by both GATK and VarScan2 were considered for further analyses.

#### Annotation of genetic variants

Genetic variants were annotated based on their prevalence in the human population, functional consequences and deleteriousness. Considering that the HSAM phenomenon is extremely rare, we retained variants absent from gnomAD v2.1.1, which comprises 125,748 exomes and 15,708 genomes^65^. This filter restricted the initial experimental selection, but was not interpreted as evidence excluding contributions from variants present in the reference dataset. Furthermore, we filtered out variants with any carriers in 253 unrelated control individuals sequenced together with the HSAM trio, or any carriers in unrelated control individuals of European ancestry from six additional WES batches of our exome studies (*N_batch1_* = 94; *N_batch2_* = 94; *N_batch3_* = 376; *N_batch4_* = 282; *N_batch5_* = 187; *N_batch6_* = 285), processed in the same manner as the WES batch including the HSAM participant. We took this step to exclude false-positive variants due to potential sequencing or data processing noise, specific to our data and processing pipeline.

The impact of variants on genes was determined using the Ensembl Variant Effect Predictor (VEP) ^66^. Variants with high impact (splice acceptor variant, splice donor variant, stop gained, frameshift variant, stop lost, start lost) or moderate impact (inframe insertion, inframe deletion, missense variant, protein altering variant) on protein coding genes, as defined by VEP (https://www.ensembl.org/info/genome/variation/prediction/predicted_data.html), were considered of interest. In cases where one variant was assigned to multiple genes, the functional consequences for each gene were considered separately. If a variant had multiple functional consequences for a single gene, the functional consequences with highest impact were taken for annotation. Predicted variant deleteriousness was additionally annotated using PHRED-scaled Combined Annotation Dependent Depletion (CADD) scores from version 1.4 for GRCh37/hg19.

#### Sanger sequencing of the locus harboring the *de novo MYCBP2* variant in the trio

The *MYCBP2* locus was amplified using the primer pairs 5’-TCATTGTCTCTTACCCTCCGG-3’/5’-TGGGAGGCTTTCTTTGGTCT-3’ and 5’-TGCAAAGAAATGTGTTCTGCA-3’/5’-AGCAATGGATGAAGACCTGGA-3’ via polymerase chain reaction (PCR) employing the KAPA HiFi HotStart PCR Kit (standard Protocol Mix, KK2501). The PCR conditions were as follows: initial denaturation at 95°C for 3 min, followed by 35 cycles of denaturation at 95°C for 30 seconds, annealing at 60°C for 30 seconds, and extension at 72°C for 1 min, with a final extension at 72°C for 5 min and a hold at 4°C. Following amplification, the PCR products were excised from the gel and purified using the Zymoclean Gel DNA Recovery Kit (D4001). The purified gel fragments were subsequently cloned into the pGEM®-T Easy Vector System (A1360) using the standard 10 µl ligation protocol. The ligation mix was used to transform JM109 High Efficiency Competent Cells under selection conditions (LB medium supplemented with ampicillin, IPTG, and X-Gal). Colonies presenting with white coloration on X-Gal plates were selected and cultured overnight in 5 ml of LB medium containing ampicillin. Plasmid DNA was then extracted from these cultures using the ZymoPURE Plasmid Miniprep Kit (D4208T). Finally, the extracted plasmids were Sanger sequenced using the standard T7 primer (TAATACGACTCACTATAGGG). Sequencing was conducted at Microsynth.

#### Long-read whole-genome sequencing and structural variant prioritization

Long-read whole-genome sequencing was performed on JN’s genomic DNA by Novogene (Novogene GmbH, Munich, Germany) using PacBio HiFi sequencing on a PacBio Revio instrument. Whole blood was collected into 10-mL BD Vacutainer Plus tubes containing spray-coated K₂EDTA (Becton Dickinson). Genomic DNA was isolated using the QIAamp DNA Blood Maxi Kit (QIAGEN, Hilden, Germany), following the manufacturer’s recommended spin protocol. Before library preparation, DNA was quantified using a Qubit fluorometer and assessed by NanoDrop spectrophotometry, conventional agarose gel electrophoresis and pulsed-field gel electrophoresis. The submitted sample contained 19.05 µg DNA at 107 ng/µl, with A260/A280 and A260/A230 ratios of 1.87 and 2.253, respectively. Electrophoresis showed a main genomic DNA band ≥30 kb. PacBio Revio sequencing generated 7,378,920 HiFi reads comprising 148.20 Gb, with a mean read length of 20,084 bp and a read-length N50 of 20,279 bp. HiFi reads were aligned to the GRCh38 reference genome using pbmm2 (v1.13.1) with the CCS preset. Structural-variant (SV) calls ≥50 bp were called using Sniffles2 (v.2.3.3)^67^ with default parameters and enabled strand-based quality control. SV calls not marked as PASS or with homozygous reference genotype were excluded and retained SVs were annotated using ANNOVAR^68^.

The downstream analysis was designed as an exploratory prioritization of SV calls with potential direct gene-level consequences rather than as a comprehensive assessment of *de novo* structural variation. For non-breakend (BND) calls, we prioritized calls that overlapped annotated exonic regions and showed less than 50% reciprocal overlap with SVs in the selected population-reference callsets: CoLoRSdb (CoLoRSdb.GRCh38.v1.2.0.pbsv.jasmine.vcf.gz), the Audano et al. callset (EEE_SV-Pop_1.ALL.sites.20181204.vcf.gz)^69^, and the Human Pangenome Reference Consortium callset (hprc.GRCh38.pbsv.vcf.gz). Specifically, we prioritized SVs that (i) overlapped exonic regions and (ii) showed less than 50% reciprocal overlap with any SV reported in the population reference datasets, irrespective of variant class. Limited overlap with these reference callsets was used solely as a prioritization criterion and was not interpreted as evidence of de novo occurrence. This procedure yielded 29 candidate non-BND SV calls. Because BND calls are defined by paired breakpoints and were not directly amenable to the interval-based reciprocal-overlap procedure used for the other SV classes, they were assessed separately. BND breakpoint loci were annotated using ANNOVAR and screened for overlap with annotated exonic or splice regions. This identified one additional candidate BND call, resulting in a total of 30 candidate SV calls for trio-WES evaluation.

As part of the prioritization workflow, the proband and parental short-read WES reads were additionally aligned to GRCh38 to enable direct comparison with the long-read SV calls. Candidate loci were then inspected in the WES alignments of JN and both parents using IGV^70^. Depending on SV class and local coverage, the assessment considered sequencing depth and breakpoint-associated alignment features, including split or soft-clipped reads, local mismatches, discordant read pairs, insert-size deviations and mate-mapping positions. Evidence was compared across the trio to determine whether a reliable pattern specific to the proband was present. Regions with insufficient or non-informative WES coverage were retained as unresolved; absence of variant-supporting reads under such conditions was not interpreted as evidence against de novo occurrence.

For deletion and duplication calls, the visual assessment was complemented by exome-based copy-number analysis using the GATK GermlineCNVCaller workflow in cohort mode. The cohort comprised 150 exomes generated in the same technical batch: JN, both parents and 147 unrelated control individuals. CNV calls were retained at the following GATK quality-score thresholds: QS >50 for duplications, QS >100 for heterozygous deletions and QS >400 for homozygous deletions. Denoised copy-ratio profiles were additionally examined across the genomic interval corresponding to each long-read deletion or duplication in JN and both parents. The available alignment, CNV-call and normalized copy-ratio evidence was combined to assign each candidate SV call to one of three prioritization categories: (1) proband-specific supportive evidence, defined as alignment and/or copy-number evidence concordant with the long-read call in JN and absent in both parents despite informative coverage; (2) unresolved or not corroborated by proband-specific trio-WES evidence, including calls with insufficient coverage, non-discriminatory patterns or no reliable proband-specific evidence; and (3) evidence suggestive of a shared event, defined as concordant alignment and/or copy-number patterns in JN and at least one parent. These categories were intended to guide prioritization and were not considered definitive assignments of inheritance. The evidence underlying each classification is reported in Supplementary Table 4, with the corresponding IGV and copy-ratio displays provided in Supplementary Data 1.

#### *C. elegans* experiments

##### Strains

The *C. elegans* Bristol strain, variety N2, was used as the WT control. The following strain with an integrated array was used: *muIs32[mec-7p*::GFP]. The following CRISPR-engineered strains were used: *rpm-1(utr86[V682F])*, *rpm-1(utr93[lf])* and *rpm-1(utr112[C3535A; H3537A; H3540A*]). A list of all the strains used in this study is shown in Supplementary Table 5. All strains were outcrossed at least four times. Standard methods were used for maintaining and manipulating *C. elegans*^71^. Animals were grown at either 20°C or 25°C. The necessity to shift animals to 25°C was due to the apparent temperature-sensitive phenotype caused by the *V682F* substitution.

##### CRISPR/Cas9 engineering

A DNA-based (plasmid/oligo) co-CRISPR strategy was used to obtain the *rpm-1* modifications of interest and *dpy-10* served as the co-CRISPR marker^72^. Cas9 was expressed from pDD162^73^. The Q5 Site-directed mutagenesis kit (New England Biolabs) was used to clone the sgRNA targeting sequence of interest into a version of pDD162 lacking the Cas9 sequence^73^. In this way the sgRNA concentration could be adapted independently of Cas9. Plasmids were purified using the Midiprep kit from Qiagen and PAGE-purified repair oligos were ordered from Microsynth AG (www.microsynth.com). pDD162 and sgRNA plasmids were injected at 25-50 ng/µl and DNA repair oligos at 20-40 ng/µl. A micropipette puller (Sutter Instrument) was used together with Borosilicate Glass Capillaries (World Precision Instruments, Inc.) to prepare injection needles. The injection mix was prepared freshly and 2 µl were transferred into the needle using Microloader tips (Eppendorf). The injection setup consisted of the FemtoJet 4i pump (Eppendorf), the DMi8 microscope (Leica) and a micromanipulator (Narishige). Injected worms were transferred to 6 cm plates, stored at 15°C for 2 hours and then shifted to 25°C. Three days later, animals showing the co-CRISPR phenotype (rollers) were singled and genotyped the day after for the presence of the modification of interest. The list of oligos and plasmids used to obtain and genotype the different CRISPR/Cas9 modifications is shown in Supplementary Table 5.

##### Measuring body length and motility

A mixed population of animals was grown at 25°C. L4-stage larvae were singled onto seeded 6-cm NGM plates for each strain. Sixteen hours later, animals were imaged using a Leica M50 microscope equipped with a FlexaCam C1 camera, and body length was measured in Fiji^74^. Body-length measurements were normalised to the mean body length of WT animals. To quantify motility, animals were “activated” by picking them gently to the centre of the plate using an eyelash. After two minutes of recovery, body bends were counted for one minute.

##### Behavioural assays

Chemotaxis towards different chemicals, negative olfactory conditioning, as well as long-term associative memory (LTAM) testing were performed as previously described ^33, 75, 76^ with the following adaptations: 1) Animals usually went through only one round of conditioning instead of the regular two rounds for LTAM, as this was sufficient to observe the memory retaining phenotype of *rpm-1*(*V682F*). 2) Animals were shifted to 25°C for the 24 h recovery phase after conditioning and only remained at the regular 20°C during recovery for specific control experiments. For the assays underlying Fig. 2d, chemotaxis indices were calculated separately for individual assay plates. Animals were counted at the diacetyl spot, at the control spot and outside both spots. The number of animals counted per plate ranged from 46 to 293 across the recorded conditions. Each plotted point represents a plate-level chemotaxis index.

##### Analysis of axon termination defects

ALM and PLM axon termination defects were analyzed using the *muIs32[mec-7p::*GFP] transgenic line^36^. Staged animals were grown at either 20°C or 25°C. Day-1 adults were picked into a drop of M9 / 0.2% sodium azide on a 3% agarose pad on a microscopy slide and covered with a cover slip. ALM and PLM axon termination defects were scored as normal or defective under a fluorescent dissecting microscope (Leica M165 FC). ALM axon termination was considered defective, if a short or long hook was observed^77^. PLM axon termination was considered defective, if it extended beyond the ALM cell body or ended in a hook^77^. One hundred animals per genotype and temperature were scored.

##### Statistics

Statistical analyses for *C. elegans* experiments were performed using two-sided Wilcoxon–Mann–Whitney tests, except for axon termination defects, which were analysed using chi-square tests. Axon termination defects were scored in 100 animals per genotype and temperature condition. A two-sided *P* < 0.05 was considered statistically significant; exact *P* values are provided in the Results.

##### Structural modelling

Three structural models of the RCC1-like domain (RLD) from the RPM-1 protein were generated using the SWISS-MODEL automated modelling workflow^78, 79^, based on template Protein Data Bank (PDB) structures 8gqe, 7vgg, and 1a12. The resulting models exhibited QMEANDisCo ^80, 81^ global scores of 0.59, 0.58, and 0.54, respectively, and local confidence scores of 0.67, 0.69, and 0.58 for the V682 residue. The V682F mutation was introduced for visualisation using FoldX^82^.

### Experiments in mice

#### Generation of Mycbp2-V>F mice

The Mycbp2 V768F knock-in line was generated in-house at the Center for Transgenic Models, University of Basel, by CRISPR-mediated homology-directed repair following microinjection of 2-cell embryos. The mouse Mycbp2 gene resides on chromosome 14 (103,350,847-103,584,250 on reverse strand) with mouse V768 residue corresponding to human V768 residue affected by the V>F variant identified in JN. The gRNA target sequence tgcatg**gtg**tgcaccgtgtg<u>tgg</u> (<u>tgg</u> PAM sequence underlined, V768 **gtg** codon in bold) was selected for optimal on target activity using the CRISPOR online tool^83^. DNA double strand breaks generated by the Cas9RNP were repaired using a ssDNA HDR template resulting in the desired V768F mutation. The sequence of linear ssDNA HR template used is listed below. The F (**ttt**) codon is shown bold and three additional silent mutations preventing re-cleavage of the modified allele by Cas9 are underlined: gcaatggatgaagacctggaggaagaactggatgagaaagacgagaagtccatgatgtgccctccaggcatgcacaagtggaagc tggagcagtgcatg**ttt**tgtac<u>a</u>gt<u>c</u>tgtggagactgcactggctacggcgccagctgtgttagcagtggccgtcctgacagagttcct ggagggtaggactagacttcctagg

Gene targeting was carried out by 2-cell embryo injections^84^. Four weeks old C57BL/6J female mice underwent ovulation induction by i.p. injection of 5 IU equine chorionic gonadotrophin (PMSG; Folligon–InterVet), followed by i.p. injection of 5 IU human chorionic gonadotropin (Pregnyl–Essex Chemie) 48 h later. For the recovery of zygotes, C57BL/6J females were mated with mature males of the same strain immediately after the administration of human chorionic gonadotropin. All zygotes were collected from oviducts 24 h after the human chorionic gonadotropin injection and were then freed from any remaining cumulus cells by a 1–2 min treatment of 0.1% hyaluronidase (Sigma-Aldrich) dissolved in M2 medium (Sigma-Aldrich). Mouse embryos were cultured overnight in KSOM (Millipore) medium at 37°C and 5% CO2. Embryos that reached 2-cell stage the following morning were used for microinjections.

All microinjections were performed using a microinjection system comprised of an inverted microscope equipped with Nomarski optics (Nikon), a set of micromanipulators (Narishige), a FemtoJet microinjection unit (Eppendorf) and a micro ePore cell membrane penetrator (WPI).

The injection mixture contained crRNA:tracrRNA at 20 ng/µl, HiFi Cas9 Nuclease V3 at 200 ng/µl and the single-stranded DNA HDR template at 20 ng/µl (all from Integrated DNA Technologies, IDT). The mixture was microinjected into the nuclei of both cells of two-cell embryos until 20–30% nuclear distension was observed.

Embryos that survived microinjection were transferred on the same day into the oviducts of 8–16-wk-old pseudopregnant Crl:CD1 (ICR) females (0.5 d after coitus) that had been mated with sterile genetically vasectomized males^85^ the day before embryo transfer. Pregnant females were allowed to deliver and raise their pups until weaning age and founders were identified by PCR analysis of genomic DNA obtained from biopsy samples.

The knock-in allele was verified by cloning and Sanger sequencing of PCR products from the selected founder used to establish the breeding line. Following backcrossing of this founder to WT C57BL/6 mice, F1 offspring and selected F2 descendants were genotyped by PCR followed by direct Sanger sequencing of the amplicons. Six sequence-verified F2 animals, i.e., four heterozygotes and two homozygotes, were generated by intercrossing the F1 heterozygotes PUP-030528 and PUP-030392 and transferred to Berlin to establish the experimental colony. The pedigree of the transferred animals is provided in Supplementary Data 2. In the Berlin facility, the colony was established by IVF using two F2 males. Subsequent colony expansion involved breeding within the established *Mycbp2-V>F* line and crosses with WT C57BL/6J animals maintained at the same facility. Breeding animals and mice assigned to experiments were biopsied and genotyped. Detailed lineage information and representative genotyping records are provided in Supplementary Data 2.

#### Routine welfare monitoring of the germline line

Animals underwent twice-weekly monitoring for adverse phenotypes in addition to routine facility observations. The facility’s monitoring summary documented observations of more than 100 animals across more than three generations and reported mortality, reproduction, general behaviour, feeding behaviour and body weight as similar to WT, with no constrained phenotype identified. The line was classified as “not strained” in Form D (national line number 4611).

#### SHIRPA primary observation screen, *Mycbp2-V>F mice*

A SHIRPA primary observation screen was performed in 30 mice: 10 homozygous, 10 heterozygous and 10 WT animals, with five males and five females per genotype. Heterozygous and WT mice were 11 weeks old; homozygous mice were 26 weeks old. The screen assessed morphology, general condition, locomotor and behavioural measures, sensory responses, reflexes and neurological function according to the scoring scheme provided in Supplementary Data 3. Complete individual-level data are also provided in Supplementary Data 3. Because the homozygous group differed in age from the other groups, comparisons involving homozygotes were considered descriptive.

#### Barnes Maze Test, Mycbp2-V>F mice

The Barnes maze test measures spatial learning and memory. Prior to the tests, animals were handled and habituated to the experimenters for one week. During the training phase animals are given 4 trials a day for 4 days with an ITI of 15 min between trials. For each trial the animals are placed on a table (92cm diameter), containing 20 holes (45mm diameter) spaced evenly apart along the periphery. Under one of these holes is a nest that the animal can go into. During the training trials, the animal is allowed to explore the maze for 3 min to find the nest. To encourage the animal to search for the nest light white noise is played (un-tuned radio). Once the animal has found the nest the white noise is stopped. If the animal does not find the nest in the allotted 3 min it is guided to the nest and the white noise is stopped only after the animal has entered the nest. After the animal has entered the nest, the animal is then allowed to stay in the nest for 1 minute undisturbed. During the probe trials on day 5 and day 12 animals are returned to the table containing the 20 holes with the nest removed. During these trials the animal’s behaviour is recorded/observed for 90s. All trials were recorded by the video-tracking system Any-maze and the time spent in goal area, latency to find the nest, the distance travelled to the nest, the number of errors and the sequence of holes was recorded.

<u>Barnes Maze Reversal:</u> After the last Barnes maze test, the nest is moved to a different position. Animals are then trained for 3 days using the same protocol as the first Barnes maze. On the 4th and 11th and 52nd day animals are probed as was done in the first Barnes maze.

#### Novel Object Recognition Test, Mycbp2-V>F mice

The novel object recognition (NOR) test is used to assess cognition, specifically recognition memory, in rodents. Prior to the experiment, animals are habituated to the arena (50 cm × 50 cm open field) by being allowed to explore for 10 min over two consecutive days. During the first trial (sample phase), animals are placed in the arena with two identical objects and allowed to explore for 10 min. Following the trial, animals are returned to their home cage for an intertrial interval (ITI) of 3 days. During the second trial (novel phase), animals are reintroduced to the arena, where they encounter two objects—one of which has been replaced with a novel object. The novel object replaced the left or right original object in a balanced manner. Animals are allowed to explore the objects for 5 min. All trials are recorded using a camera mounted above the arena, and the Viewer software (BIOBSERVE) is used to track and record animal behaviour. The time spent exploring each object and the number of visits to each object are quantified, and the object-recognition discrimination index is calculated.

The behavioural experiments were approved by the State Office for Health and Social Affairs (Landesamt für Gesundheit und Soziales, LAGeSo), Berlin, under licence 2021-043-G.

#### Novel Object Recognition and Object Location Tests, Mycbp2^V768F(hSyn) mice

To assess object recognition and spatial memory following introduction of the *MYCBP2* mutation into the dorsal hippocampus via prime editing, novel object recognition (NOR) and object location (OL) tests were performed as previously described ^86, 87^, with minor modifications. Behavioural testing was conducted 6–8 weeks after surgery. Prior to testing, mice were handled for 5 min per cage per day for 5 consecutive days. Before each habituation or test session, animals were single-housed for at least 30 min to minimize potential effects of acute social disruption on behaviour. Mice were habituated to the behavioural arena (45 × 45 × 40 cm) by free exploration of the empty apparatus for 5 min. All behavioural procedures were conducted under low-light conditions (∼4 lux), with additional infrared illumination to facilitate video recording. Twenty-four hours later, mice underwent the NOR familiarization phase, during which they were allowed to explore two identical objects placed equidistant from the left and right corners of the arena for 5 min. Object identity and left–right positioning were counterbalanced across animals to control for object-and side-related preferences. Memory retention was assessed by re-exposing mice to the arena after an intertrial interval (ITI) of either 20 min (short-term memory) or 24 h (long-term memory), during which one familiar object and one novel object were presented. For the object location test, mice were re-exposed to the same familiar objects used during the NOR familiarization phase, with one object displaced to a novel location, following an ITI of either 20 min or 24 h. The arena and objects were cleaned with 10% ethanol between animals. All sessions (habituation, familiarization and test) were video recorded. Short-and long-term memory tests were performed in the same animals, separated by approximately 4 weeks. Behaviour during habituation was analysed using in-house–developed YOLO-based tracking software. Familiarization and test sessions were scored offline by two observers blinded to treatment conditions, quantifying time spent exploring each object or location as well as total exploration time. Videos exceeding a predefined variance threshold were additionally scored by a third independent blinded observer. Inter-rater reliability was high (r(307) = 0.947, *P* < 0.0001).

#### Behavioural Analysis and Statistics

All tests were two-tailed with α = 0.05. Across all studies, the analytical approach for each behavioural paradigm was defined *a priori* based on the distributional properties of the dependent variables. Normality was evaluated using Shapiro-Wilk tests. Where normality assumptions were met, parametric models were used; where they were violated, non-parametric approaches were applied as detailed below. Outlier detection for dependent variables was performed using standardized model residuals (|z| > 3). For total exploration time (TET), which served as a task-engagement metric, outlier detection was performed on standardized raw values calculated across genotype groups (|z| > 3), as extreme values may reflect abnormal exploration behaviour unrelated to memory performance. In both cases, exclusions were applied only to the respective variable and sensitivity analyses including excluded values were conducted. This procedure was defined *a priori*, applied uniformly across genotype groups and behavioural paradigms, and did not involve exclusion of entire animals across tests. Sample sizes, employed statistical tests, and *P* values for each analysis are reported in the figure legends or Results section. Behavioural assessments were conducted blinded to genotype. ANOVA and ANCOVA were conducted using SPSS (IBM SPSS Statistics, version 31). Rank-based analyses were performed in R using ARTool for aligned rank transform ANOVA^88^ and nparLD for Brunner–Langer-type nonparametric longitudinal analyses^89^. Effect sizes are reported as partial eta-squared (*η*^2^).

NOR and OL performance were quantified using the discrimination index (DI) or raw exploration differences, as specified below. The DI was calculated as the difference in time spent exploring the novel versus the familiar object (NOR), or the novel versus the previous location (OL), divided by the total exploration time during the test phase (i.e., DI = (T_novel − T_familiar) / (T_novel + T_familiar)). Total exploration time (TET) was defined as the sum of time spent exploring the two objects or locations during the test phase (i.e., TET = T_novel + T_familiar). Animals with a TET < 20 sec were *a priori* excluded from the respective analyses^87^. DI was used as it is the most reported summary measure in NOR/OL studies. Because ratio-based measures, such as DI, may produce misleading inferences under substantial mathematical coupling, covariate-adjusted regression methods such as ANCOVA are often preferable^90, 91^. Therefore, the association between DI and TET was evaluated using Spearman’s rank correlation (ρ) prior to subsequent analyses, and an *a priori* effect-size criterion was applied. When DI showed limited dependence on TET (|ρ| < 0.30, corresponding to less than a moderate effect size), genotype effects were tested using linear models with DI as the dependent variable. When DI showed relevant substantial coupling to TET (|ρ| ≥ 0.30), genotype effects were assessed using analysis of covariance (ANCOVA) on the raw exploration difference score (ΔT = T_novel − T_familiar), with TET included as a covariate.

In all models, genotype, sex and cohort (two experimental runs) were included as fixed factors, and genotype × sex interactions were evaluated to account for potential sex-dependent effects, in accordance with current recommendations for investigating sex-dependent effects in biomedical research. Residuals from the linear models (ANOVA or ANCOVA, as appropriate) were checked for normality using the Shapiro-Wilk test.

To determine whether animals exhibited memory for the novel object or location during the probe trials, within-genotype comparisons of exploration time directed toward the novel versus familiar object or location were performed using paired-samples t-tests or Wilcoxon signed-rank tests, as appropriate.

For experiments including short (20 min) and long (24 h) inter-trial intervals (ITI), analyses were conducted separately for each delay condition, and between-genotype comparisons were performed within each ITI.

For the Barnes maze, the primary outcome measure was time spent in the target zone during probe trials. The models for the initial and reversal phases included probe (1d and 8d for initial; 1d, 8d, 49d for reversal) as a within-subject factor and genotype, sex, and cohort (two experimental runs) as between-subject factors. Residuals from the repeated-measures ANOVA model of the initial phase fulfilled normality assumptions (Shapiro-Wilk *P* > 0.11 across both probes). Residuals from the repeated-measures ANOVA model of the reversal phase deviated significantly from normality (Shapiro-Wilk *P* < 0.001 across all probes), and inspection of residual distributions indicated substantial deviation from Gaussian assumptions that could not be corrected by transformation. Hence, the primary inference was based on the rank-based Aligned Rank Transform (ART) ANOVA ^88^ using ARTool (https://depts.washington.edu/acelab/proj/art/). As a sensitivity analysis, we performed a Brunner-Langer-type rank-based longitudinal analysis (nparLD framework) and report the ANOVA-type statistic (mATS) for the main effect of genotype. As nparLD allows only two between-subject factors, we modelled sex and cohort as a combined factor and included this composite factor alongside genotype, with probe as the within-subject factor.

#### Prime editing

Prime editors (PEs) are programmable gene-writing systems in which a Cas9 nickase directs target recognition while a tethered reverse transcriptase copies an edit encoded in a prime-editing guide (peg)RNA directly into the genome ^92–96^. To introduce the *MYCBP2* variant into adult mice, we first screened nine pegRNA designs ^97^ encoding the GTG-to-TTC edit at the *Mycbp2* locus in mammalian cell lines (Supplementary Fig. 3a). pegRNA vectors were co-transfected with a PE-expressing plasmid into murine Hepa1-6 and human K562 reporter cells carrying a stably integrated murine *Mycbp2* target site (hereafter referred to as Hepa and K562), followed by analysis of editing rates via deep amplicon sequencing at 7 days post-transfection (Supplementary Fig. 3b). pegRNA3 showed highest on-target editing efficiency (Hepa, 10.6±4.0 %; K562, 14.4±9.9%; Supplementary Fig. 3c,d) without generating indels above baseline (Hepa,0.59±0.13 %; K562, 0.39±0.15%; Supplementary Fig. 3c,d), and was therefore selected for in vivo application. For neuronal delivery, we packaged an intein-split PE expressed from the hSyn1 promoter together with pegRNA3 into two AAV9 vectors, with the pegRNA cassette encoded on the N-terminal PE vector (Fig. 3c). Using stereotactic injection of AAV9-hsyn1-EGFP particles into adult mice as a surrogate (Supplementary Fig. 4a), we first confirmed efficient (Supplementary Fig. 4b,c) and neuron-specific viral spread (Supplementary Fig. 4d) throughout the dHC, including regions CA3, CA4, and the dentate gyrus (DG; Supplementary Fig. 4c), regions found to be enlarged in JN (Fig. 1b,c; Supplementary Fig. 1). Next, we delivered N-and C-terminal PE-expressing AAV particles bilaterally to the dHC of adult mice by stereotactic injection at a dose of 1 × 10⁹ vector genomes (vg) per animal (Fig. 3d). Control animals received AAV9 particles encoding a non-targeted PE lacking the pegRNA3 expression cassette. After a 6-week recovery period, we assessed whether the installed *MYCBP2* variant induces superior long-term memory similar to our previous observations (Fig. 3e).

#### Prime editing: generation of plasmids

To generate pegRNA plasmids, annealed spacer, scaffold, and 3’ extension oligos were cloned into the BsaI-digested pU6-tevopreQ1-GG acceptor plasmid (Addgene #174038) using Golden Gate assembly as previously described^97^. To generate intein-split PE plasmids, inserts were amplified from plasmids using PCR and assembled using HiFi DNA Assembly Master Mix [New England Biolabs (NEB)]^94^. To generate PiggyBac transposon plasmids for the *Mycbp2* target site, inserts with homology overhangs were cloned into the XbaI-and EcoRI-digested pPB-Zeocin backbone using HiFi DNA Assembly Master Mix (NEB). To prepare plasmids for AAV production, inserts with homology overhangs were generated by PCR and cloned into XbaI-and NotI-digested AAV backbones using HiFi DNA Assembly Master Mix (NEB). All PCRs were performed using Q5 High-Fidelity DNA Polymerase (NEB). The identity of all plasmids was confirmed by long-read sequencing. Primers used for cloning of all plasmids are listed in Supplementary Table 6.

#### Prime editing: cell culture transfection and genomic DNA preparation

Hepa1-6 (ATCC CRL-1830) cells were maintained in Dulbecco’s modified Eagle’s medium (DMEM) plus GlutaMAX (Thermo Fisher Scientific), supplemented with 10% (v/v) fetal bovine serum (FBS; Sigma-Aldrich) and 1% penicillin/streptomycin (Thermo Fisher Scientific) at 37°C and 5% CO_2_. K562 cells (ATCC CCL-243) were maintained in RPMI medium plus GlutaMAX (Thermo Fisher Scientific), supplemented with 10% (v/v) FBS and 1% penicillin/streptomycin (Thermo Fisher Scientific). Cells were passaged every 3 to 4 days and maintained at confluency below 90%. All cell lines tested negative for Mycoplasma contamination.

Cells were seeded in 96-well cell culture plates (Greiner) and transfected at 70% confluency using 0.5 μl Lipofectamine^TM^ 2000 (Thermo Fisher Scientific). If not stated otherwise, 300 ng of PE and 100 ng of pegRNA were used for transfection. Cells were incubated for 7 days after transfection. Genomic DNA from cells was isolated using a lysis buffer (10 mM Tris-HCl, 2 % Triton™ X-100, 1 mM EDTA, proteinase K [20 mg/mL]; Thermo Fisher Scientific). Cells were lysed at 60°C for 1 h, followed by 10 min at 95°C. Lysates were further diluted with 60 µL of dH_2_O. Genomic DNA from mouse tissues was isolated by phenol/chloroform extraction. First, tissue samples were incubated overnight in lysis buffer (50 mM Tris-HCl pH 8.0, 100 mM EDTA, 100 mM sodium chloride, and 1% SDS; Thermo Fisher Scientific) at 55°C and 300 rpm. Subsequently, phenol/chloroform/isoamyl alcohol (25:24:1, Thermo Fisher Scientific) was added and samples were centrifuged (5 min, maximum speed). The upper phase was transferred to a clean tube and DNA was precipitated using 100% ethanol (Sigma-Aldrich). Samples were centrifuged (5 min, maximum speed) and pellets were washed using cold 70% ethanol (−20°C). Washed pellets were dried at 55°C for 10 min and resuspended in 100 µL of dH2O.

#### Prime editing: generation of cell lines with the integrated Mycbp2 target site

To generate Mycbp2-V768F cell lines with the PiggyBac transposon, Hepa1-6 and K562 cells were seeded into a 48-well cell culture plate (Greiner) and transfected at 70% confluency with 225 ng of the PiggyBac-transposon and 25 ng of the transposase using Lipofectamine 2000 (Thermo Fisher Scientific) according to the manufacturer’s instructions. Three days after transfection, cells were enriched for 10 days using Zeocin selection [150 μg/ml].

#### Prime editing: AAV production

For the production of pseudo-typed AAV9 vectors (AAV2/9), packaging, capsid, and helper plasmids (Addgene no. 112865 and no. 112867, a gift from James M. Wilson) were co-transfected in HEK293T cells and incubated for six days until harvest. AAV particles were precipitated using PEG (VWR) and NaCl (Sigma-Aldrich) and subjected to gradient centrifugation with OptiPrep (Sigma-Aldrich) for further purification. Subsequently, the concentrated vectors were obtained using Vivaspin® 20 centrifugal concentrators (VWR). Physical titers (vector genomes per milliliter, vg/mL) were measured using a Qubit 3.0 fluorometer and the dsDNA HS assay kit (Thermo Fisher Scientific). AAV2/9 viruses were stored at −80°C until used. All other pseudo-typed AAV2/9 vectors were produced by the Viral Vector Facility of the Neuroscience Center Zurich.

#### Prime editing: amplification for deep sequencing

Genomic DNA from cultured cells or brain tissues was isolated by direct lysis (cells) or phenol/chloroform extraction (brain tissue).

*Mycbp2*-specific primers were used to generate targeted amplicons for deep sequencing. Input genomic DNA was first amplified in a 10μL reaction for 25 cycles using NEBNext High-Fidelity 2×PCR Master Mix (NEB). Amplicons were purified using AMPure XP beads (Beckman Coulter) and subsequently amplified for eight cycles using primers with sequencing adapters. Approximately equal amounts of PCR products were pooled, gel purified, and quantified using a Qubit 3.0 fluorometer and the dsDNA HS Assay Kit (Thermo Fisher Scientific). Paired-end sequencing of purified libraries was performed on an Illumina Miseq. Primers for deep sequencing are listed in Supplementary Table 7.

#### Prime editing: HTS data analysis

Sequencing reads were first demultiplexed using the Miseq Reporter (Illumina). Next, amplicon sequences were aligned to their reference sequences using CRISPResso2 ^98^. Prime editing efficiencies were calculated as percentage of (number of reads containing only the desired edit)/(number of total aligned reads). Indel rates were calculated as percentage of (number of indel-containing reads)/(total aligned reads).

#### Prime editing: statistical analysis and reproducibility

All statistical analyses were performed using GraphPad Prism (version 10.2.1). If not stated otherwise, data are represented as biological replicates and are depicted as means±standard deviation (s.d.). Statistical analyses are indicated in the corresponding figure legends. Likewise, sample sizes and the statistical tests performed are described in the respective figure legends. The data were tested for normality using the Shapiro-Wilk test if not stated otherwise. For all analyses, *P*<0.05 was considered statistically significant. For analysis of editing efficiencies, researchers were not blinded to group allocation. Blinding was not necessary here because the editing efficiency as a readout cannot be influenced by a biased researcher. All deep amplicon sequencing data was analysed via an unblinded operator by using an automated script (CRISPResso2) with limited experimenter intervention. For performance and analysis of behaviour, researchers were blinded throughout the study. Biological replicates, animal numbers, and group sizes are indicated for each experiment in the respective figure legends.

#### Stereotactic surgery

C57BL/6JRj DBH-Cre heterozygous mice (9–13 weeks old) were subjected to stereotactic surgery for delivery of prime-editing constructs; behavioral testing commenced 6–8 weeks after surgery, when mice were approximately 16–21 weeks old, with the later behavioral experiments conducted at approximately 20–25 weeks of age. Anesthesia was induced with 4% isoflurane and maintained at 2% isoflurane throughout the procedure using an automated stereotaxic instrument with automated nanoinjector (RWD #71001-S/71001-D). Analgesia consisted of subcutaneous meloxicam (5 mg kg⁻¹) and buprenorphine (0.1 mg kg⁻¹), with local application of lidocaine and bupivacaine (each 2 mg kg⁻¹) before and after surgery. After skull exposure, bregma and lambda were identified as the intersections of the coronal/sagittal and sagittal/lambdoid sutures, respectively, and skull alignment was automatically corrected for tilt and scaling using stereotaxic software (RWD). Bilateral craniotomies were made above the dorsal hippocampus at −1.7 mm anteroposterior (AP) and ±1.5 mm mediolateral (ML) relative to bregma. Mice received bilateral injections of either *phSyn-Pemax(1–713)-pegRNA3-Mycbp2* (aavSL 125) or a non-targeting Pemax control (aavSL 130-v1.1) using a nanoinjection pump (RWD). Per hemisphere, two injections of 0.5 µl each (titre 1 × 10¹² vg ml⁻¹; total dose 1 × 10⁹ vg) were delivered at dorsoventral depths of −2.3 mm and −1.5 mm at a rate of 23 nl s⁻¹. After the first injection, the capillary was raised to the second depth to allow continuous viral deposition along the longitudinal axis of the dorsal hippocampus. After the second injection, the capillary was left in place for 5 min before withdrawal. On-target prime editing at the *Mycbp2* locus was confirmed in all treated animals with editing efficiencies reaching up to 54.0% (Supplementary Fig. 3e). Because editing efficiencies were determined by deep sequencing of bulk dorsal hippocampal DNA, these measurements quantify edited alleles but do not resolve whether individual edited cells carried monoallelic or biallelic edits.

#### Histological verification of viral expression, Mycbp2^V768F(hSyn) mice

Histology was performed to verify viral spread within the dorsal hippocampus and neuron - specific transgene expression driven by the hSyn promoter as described previously ^99^. In a separate reporter-virus cohort, viral spread and neuron-specific expression were assessed using an AAV construct of the same serotype and driven by the same neuronal hSyn promoter as the prime-editing vectors, but expressing EGFP instead of the MYCBP2 prime editor (ssAAV9/2-hSyn1-EGFP-WPRE-hGHp(A)). The reporter virus was delivered using the same stereotactic injection procedure to adult mice aged 15–16 weeks. Brains were collected approximately 5 weeks later, at 20–21 weeks of age, to assess viral distribution and neuronal expression throughout the dorsal hippocampus. Approximately 5 weeks after injection, mice were deeply anesthetized with pentobarbital (150 mg kg⁻¹, i.p.) and transcardially perfused with phosphate-buffered saline (PBS) followed by 4% paraformaldehyde (PFA). Brains were extracted, post-fixed in 4% PFA for 4 h, and cryoprotected in 30% sucrose in PBS. Coronal brain sections (40 µm) were prepared using a cryostat and processed as free-floating sections. Sections were incubated overnight at 4 °C with a primary antibody against NeuN (rabbit anti-NeuN; Abcam ab177487; 1:400) diluted in PBS containing 4% normal serum and 0.05% Triton X-100. After washing, sections were incubated for 1 h with an Alexa Fluor 647– conjugated secondary antibody (goat anti-rabbit; Invitrogen A32733; 1:500). Sections were mounted with an aqueous medium containing DAPI. Fluorescence images were acquired using a Zeiss Axio Observer widefield microscope and a Zeiss LSM 880 confocal microscope.

## Notes

### Competing Interest Statement

The authors have declared no competing interest.

### Author Declarations

- The study protocol was approved by the Ethics Committee of Northwest and Central Switzerland. - The behavioural experiments were approved by the State Office for Health and Social Affairs (Landesamt fur Gesundheit und Soziales (LAGeSo), Berlin, under licence 2021-043-G. - Data for the control population were obtained from the UK Biobank through research application number 101531. Informed consent was obtained by the UK Biobank for all participants.

### Summary of Updates

Methods, Results, and Discussion updated to clarify. Figure 1, Supplementary Figures, and Supplementary Tables 3 and 4 updated to clarify.

