## Supplemental Figures for "A *de novo* human *MYCBP2* variant enhances memory persistence across species"

### Supplementary figures

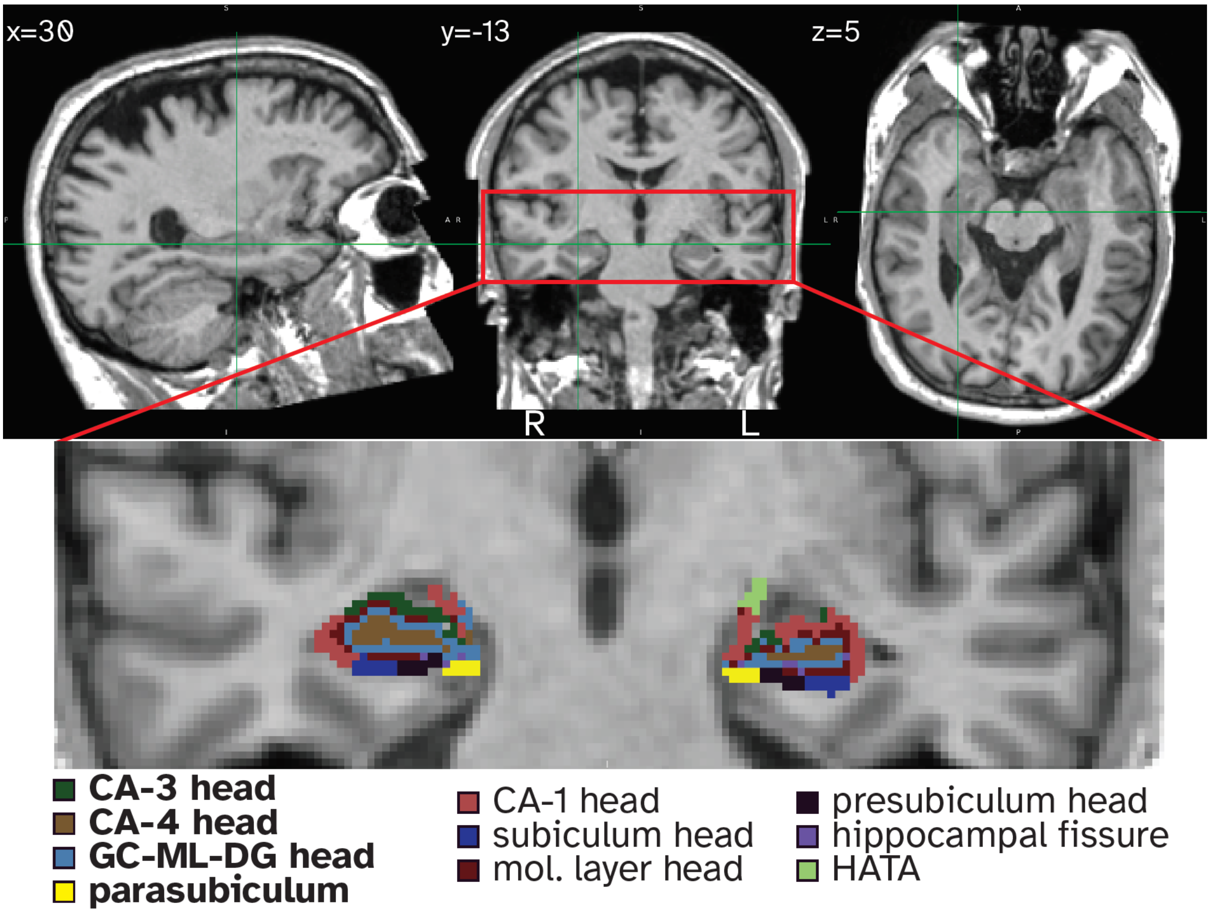


**Supplementary Fig. 1 | Structural MRI and hippocampal subfield segmentation in the HSAM participant.** T1-weighted structural MRI of JN shown in sagittal (left), coronal (middle), and axial (right) views. Crosshairs indicate the position of the displayed coronal section (y = −13), which is enlarged below. The enlarged coronal image shows the FreeSurfer v6.0 hippocampal subfield segmentation overlaid on the structural MRI. The four right hippocampal subfields with volumes exceeding Z = 3 relative to the age- and sex-matched UK Biobank reference sample—CA3 head, CA4 head, GC-ML-DG head, and parasubiculum—are indicated in bold in the legend. GC-ML-DG, granule cell and molecular layers of the dentate gyrus; HATA, hippocampal-amygdaloid transition area; R, right; L, left.

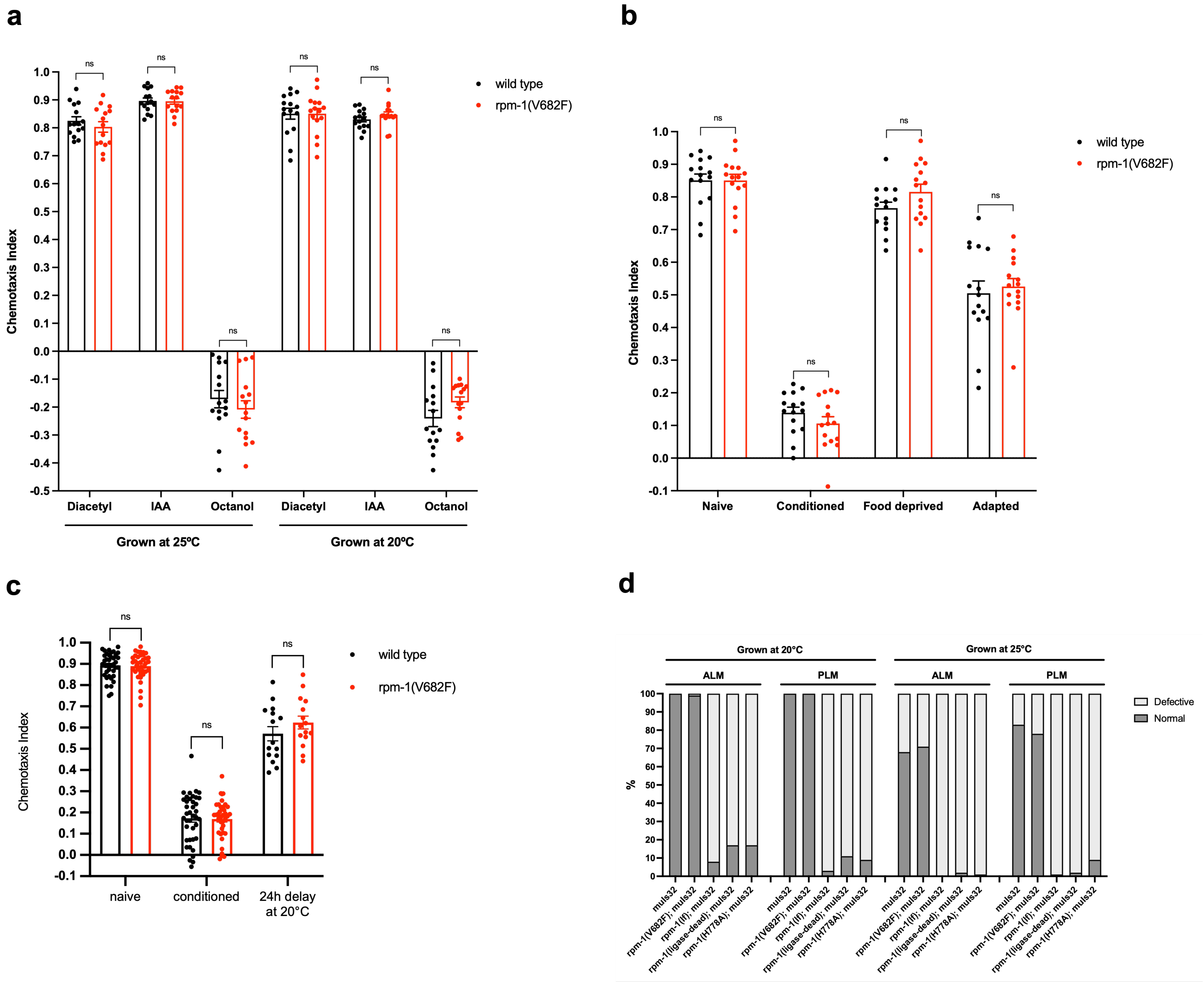


**Supplementary Fig. 2 | Impact of the mutated residue on sensory, behavioral, and axonal measurements in *C. elegans*. a**, Chemotaxis of naïve adult N2 WT and *rpm-1* mutant worms (*rpm-1(V682F)*) towards two volatile attractants (diacetyl (DA), isoamyl alcohol (IAA)) and a repellent (octanol) under two different growth conditions (i.e., 25°C and 20°C). Dots represent single plates. **b**, Chemotaxis of adult N2 WT and *rpm-1* mutant worms (*rpm-1(V682F)*) towards DA under four experimental conditions: naïve (unconditioned), conditioned (directly after 1h of food deprivation in the presence of DA), food deprived (worms placed on a plate devoid of both food and DA for a duration of one hour), adapted (worms placed on a plate containing both food and DA for a duration of one hour). Dots represent single plates. **c**, Chemotaxis indices towards DA of WT and *rpm-1(V682F)* worms under three conditions: naïve (unconditioned), conditioned (directly after 1h of food deprivation in the presence of DA), and 24h after conditioning. For the last condition, worms were kept at 20°C during 24h after conditioning. Dots represent single plates. For bar plots **a-c**, bars and error bars represent means and s.e.m., respectively. All *P* values related to these plots are two-sided (non-parametric Wilcoxon-Mann-Whitney test), ns: $P \geq 0.05$. **d**, Quantification of axon termination in the anterior lateral mechanosensory (ALM) and posterior lateral mechanosensory (PLM) neurons utilizing a strain that expresses GFP in these neurons (*muIs32 [mec-7p::GFP]*). Worms were grown under two different temperature conditions. Conventional *rpm-1* loss-of function mutants (*rpm-1(lf)*) and animals harboring variants that severely affect RPM-1 ubiquitin ligase activity (*rpm-1(LD)*) display severe axon termination defects. Axon termination in *rpm-1(V682F)* mutants is not impaired under physiological growing conditions and is comparable to that of N2 animals under conditions known to affect axon development (worms grown at 25°C). Each bar represents analysis of axon termination in 100 worms. Light gray bars represent worms that exhibited defective axon termination.

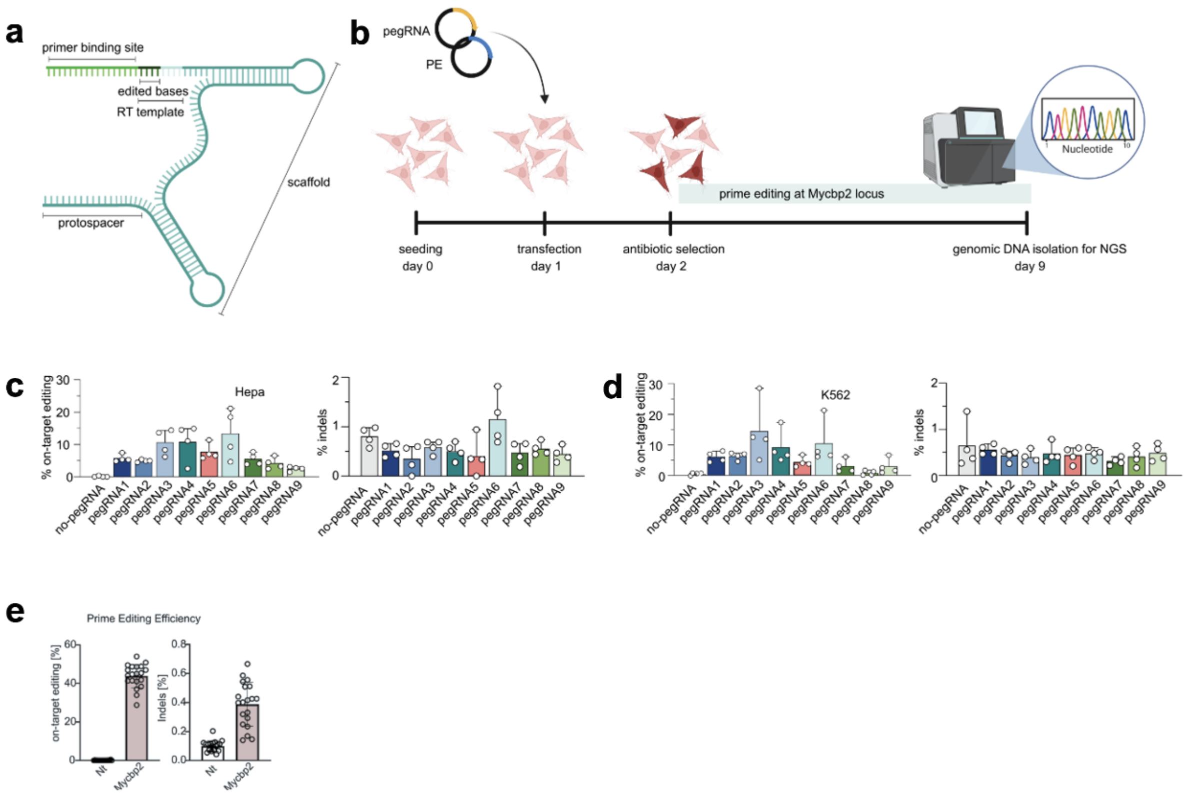


**Supplementary Fig. 3 | In vitro screening of pegRNA activity at the *Mycbp2* locus in cell lines and prime editing efficiency in treated animals. a,** Schematic representation of the components of a pegRNA, including the protospacer, the RT template encoding the desired edit, and the primer binding site, priming reverse transcription. **b,** Schematic representation of the experimental setup and timeline to assess pegRNA activity in cell lines. Created in BioRender (https://BioRender.com). **c,d,** On-target editing (left) and indel rates (right) of nine pegRNAs in Hepa **(c)** and K562 cells **(d)** (*n* = 4 biological replicates). **e,** On-target editing and indel rates at the *Mycbp2* locus in the dorsal hippocampus (dHC) of treated mice at 23-31 weeks post-injection. Animals were treated with AAV9-PEmax-nT Mycbp2 or a non-targeting control (Nt).

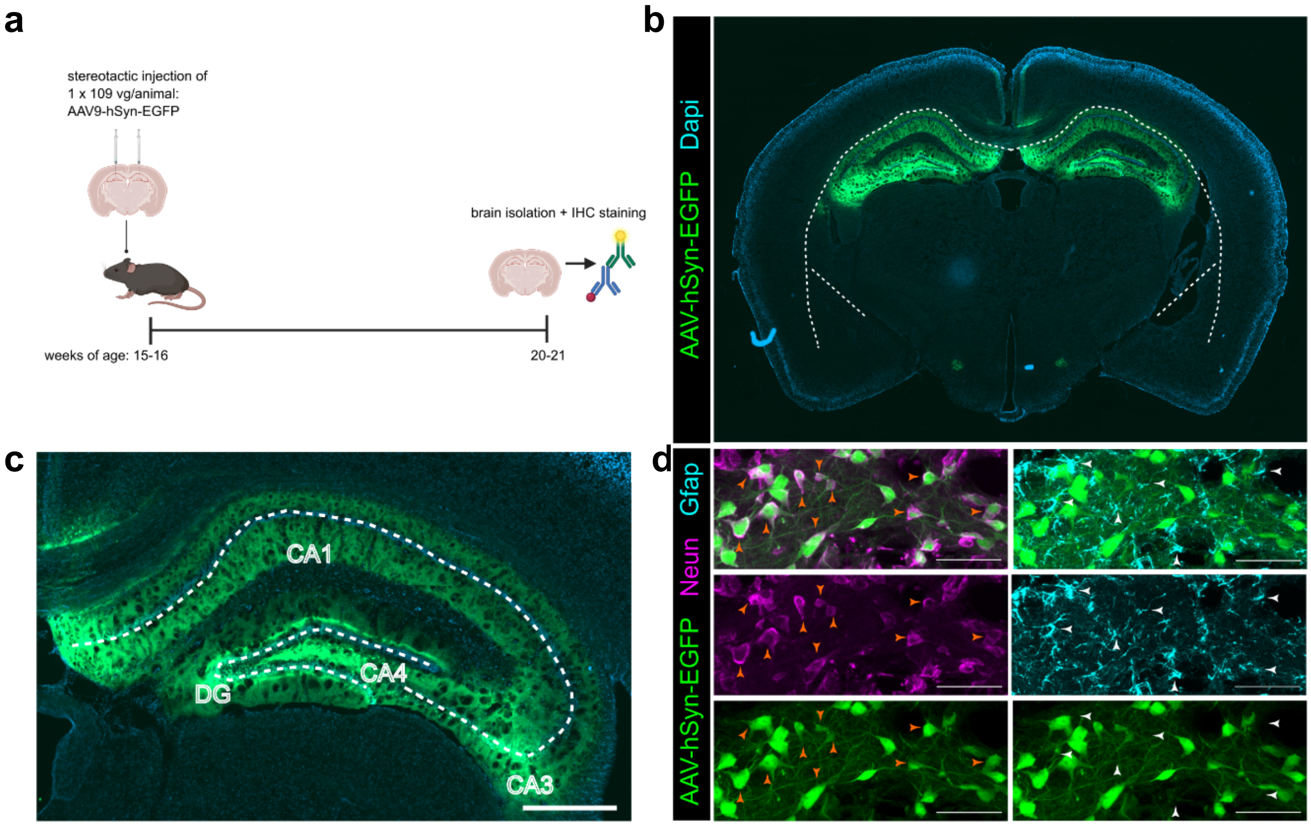


**Supplementary Fig. 4 | Verification of viral spread and neuron-specific expression for in vivo MYCBP2 prime editing. a,** Schematic representation of the experimental design and timeline. Created in BioRender (https://BioRender.com). **b,** Coronal section (40 µm) of mouse hippocampus AAV-hSyn-EGFP in green, nuclei stained with DAPI in blue. **c,** Graphical depiction of dorsal hippocampal subregions at injection site from **b**, scale bar 500 µm. **d,** Neun positive neurons (magenta), but not Gfap positive astrocytes (blue), overlay with EGFP (green); scale bars 50 µm.
