## Supplemental Table 1 for "A *de novo* human *MYCBP2* variant enhances memory persistence across species"

**Supplementary Table 1.** Detailed neuropsychological test results.

| Assessment  Battery | Subtest | Raw Score | Scaled or t-score | Transformed score | Percentile | Level of Ability |
| --- | --- | --- | --- | --- | --- | --- |
| WAIS-IV |  |  |  |  |  |  |
| *subtests* | Block design | 38 | 11^s^ |  |  |  |
|  | Similarities | 28 | 11^s^ |  |  |  |
|  | Digit Span | 34 | 14^s^ |  |  |  |
|  | Matrix Reasoning | 18 | 12^s^ |  |  |  |
|  | Vocabulary | 49 | 13^s^ |  |  |  |
|  | Arithmetic | 15 | 11^s^ |  |  |  |
|  | Symbol Search | 27 | 10^s^ |  |  |  |
|  | Visual Puzzles | 7 | 06^s^ |  |  |  |
|  | Information | 14 | 10^s^ |  |  |  |
|  | Coding | 55 | 09^s^ |  |  |  |
| *domains* | Verbal Comprehension | - | - | 107 (101-112) | 68 | average |
|  | Perceptual Reasoning | - | - | 98 (92-104) | 45 | average |
|  | Working Memory | - | - | 114 (106-120) | 82 | high average |
|  | Processing Speed | - | - | 97 (89-106) | 42 | average |
|  | Full-Scale IQ (FSIQ) | - | - | 104 (100-108) | 61 | average |
| WMS-IV |  |  |  |  |  |  |
| *subtests* | Logical memory 1 | 32 | 13^s^ |  |  |  |
|  | Logical memory 2 | 25 | 12^s^ |  |  |  |
|  | Verbal paired associates 1 | 29 | 10^s^ |  |  |  |
|  | Verbal paired associates 2 | 10 | 11^s^ |  |  |  |
|  | Designs 1 | 62 | 09^s^ |  |  |  |
|  | Designs 2 | 50 | 10^s^ |  |  |  |
|  | Visual Reproduction 1 | 39 | 12^s^ |  |  |  |
|  | Visual Reproduction 2 | 30 | 12^s^ |  |  |  |
| *domains* | Auditory Memory | - | - | 109 (102-115) | 73 | average |
|  | Visual Memory | - | - | 104 (98-109) | 61 | average |
|  | Immediate Memory | - | - | 107 (100-113) | 68 | average |
|  | Delayed Memory | - | - | 108 (101-114) | 70 | average |
| Stroop | Word | 114 | 55^t^ |  |  | average |
|  | Color | 49 | 34^t^ |  |  | borderline |
|  | Color-Word | 35 | 45^t^ |  |  | average |
|  | Interference | 35 | 52^t^ |  |  | average |

**Raw score:** The raw score of the corresponding subtest.

**Scaled or t-score:**

s: Raw score transferred to a scaled score with an age-matched reference population. The scaled score is centered at a mean of 10 and a standard deviation of 3.

t: Raw score transferred to a T-score with an age- and education-matched reference population. The T-score is centered at a mean of 50 and a standard deviation of 10.

**Transformed Score**: Index scores and composite scores of WMS-IV were calculated for each domain from the corresponding subtests. The number represents the value and the 95% confidence interval.

**Percentile:** Percentile rank in relation to the normative population.

**Level of Ability:** The participant's level of ability in the respective domain.
