## Supplemental Table 6 for "A *de novo* human *MYCBP2* variant enhances memory persistence across species"

**Supplementary Table 6.** Primers used for cloning of all plasmids in the prime editing experiment.

| pegRNA36_Mycbp2_spacer_fwd | CACCGTGGAAGCTGGAGCAGTGCAGTTTC |
| --- | --- |
| pegRNA36_Mycbp2_spacer_rev | CTCTGAAACTGCACTGCTCCAGCTTCCAC |
| pegRNA36.1_Mycbp2_ext_fwd | GTGCAGTCTCCACACACGGTGCAgaaCATGCACTGCTCCAGCTTC |
| pegRNA36.1_Mycbp2_ext_rev | CGCGGAAGCTGGAGCAGTGCATGttcTGCACCGTGTGTGGAGACT |
| pegRNA36.2_Mycbp2_ext_fwd | GTGCAGTCTCCACACACGGTGCAgaaCATGCACTGCTCCAGCTTCC |
| pegRNA36.2_Mycbp2_ext_rev | CGCGGGAAGCTGGAGCAGTGCATGttcTGCACCGTGTGTGGAGACT |
| pegRNA36.3_Mycbp2_ext_fwd | GTGCGGTGCAgaaCATGCACTGCTCCAGCTTCC |
| pegRNA36.3_Mycbp2_ext_rev | CGCGGGAAGCTGGAGCAGTGCATGttcTGCACC |
| pegRNA36.4_Mycbp2-SM1_ext_fwd | GTGCAGTCTCCACACACGGTGCAgaacataCACTGCTCCAGCTTC |
| pegRNA36.4_Mycbp2-SM1_ext_rev | CGCGGAAGCTGGAGCAGTGtatgttcTGCACCGTGTGTGGAGACT |
| pegRNA36.5_Mycbp2-SM1_ext_fwd | GTGCAGTCTCCACACACGGTGCAgaacataCACTGCTCCAGCTTCC |
| pegRNA36.5_Mycbp2-SM1_ext_rev | CGCGGGAAGCTGGAGCAGTGtatgttcTGCACCGTGTGTGGAGACT |
| pegRNA36.6_Mycbp2-SM1_ext_fwd | GTGCAGTCTCCACACACGGTGCAgaacataCACTGCTCCAGCTT |
| pegRNA36.6_Mycbp2-SM1_ext_rev | CGCGAAGCTGGAGCAGTGtatgttcTGCACCGTGTGTGGAGACT |
| pegRNA36.7_Mycbp2-SM2_ext_fwd | GTGCAGTCTCCACACACGGTacagaaCATGCACTGCTCCAGCTTC |
| pegRNA36.7_Mycbp2-SM2_ext_rev | CGCGGAAGCTGGAGCAGTGCATGttctgtACCGTGTGTGGAGACT |
| pegRNA36.8_Mycbp2-SM2_ext_fwd | GTGCAGTCTCCACACACGGTacagaaCATGCACTGCTCCAGCTTCC |
| pegRNA36.8_Mycbp2-SM2_ext_rev | CGCGGGAAGCTGGAGCAGTGCATGttctgtACCGTGTGTGGAGACT |
| pegRNA36.9_Mycbp2-SM2_ext_fwd | GTGCAGTCTCCACACACGGTacagaaCATGCACTGCTCCAGCTT |
| pegRNA36.9_Mycbp2-SM2_ext_rev | CGCGAAGCTGGAGCAGTGCATGttctgtACCGTGTGTGGAGACT |
| pegRNA_opt-scaffold_fw | AGAGCTATGCTGGAAACAGCATAGCAAGTTGAAATAAGGCTAGTCCGTTATCAACTTGAAAAAGTGGCACCGAGTCG |
| pegRNA_opt-scaffold_rev | GCACCGACTCGGTGCCACTTTTTCAAGTTGATAACGGACTAGCCTTATTTCAACTTGCTATGCTGTTTCCAGCATAG |
| DB215_epegRNA-Mycbp2_rev | gatgcggtgggctctatggCTCGAGCGGCCCAAGCTTAAAAAAATTC |
| DB136_pegRNA-U6-NdeI_fwd | tgttttaaaatggactatcatatgCTTACCGTAACTTGAAAGTATTTC |
