## Supplemental Table 7 for "A *de novo* human *MYCBP2* variant enhances memory persistence across species"

**Supplementary Table 7.** Primers used for deep sequencing in the prime editing experiment.

| NGS_Mycbp2-endo-invivo_fwd | CTTTCCCTACACGACGCTCTTCCGATCTNNNNNNAAGTCCATGATGTGCCCTCC |
| --- | --- |
| NGS_Mycbp2-endo-invivo_rev | GGAGTTCAGACGTGTGCTCTTCCGATCTNNNNNNACCCTCCAGGAACTCTGTCA |
| NGS_Mycbp2-reporter-invivo_fwd | CTTTCCCTACACGACGCTCTTCCGATCTNNNNNNTGTGCTGGAATTCGGTTTGG |
| NGS_Mycbp2-reporter-invivo_rev | GGAGTTCAGACGTGTGCTCTTCCGATCTNNNNNNCTCTAGAGTCAGGACGGCCA |
